# No evidence for a classic transmission-duration tradeoff in human malaria infections

**DOI:** 10.64898/2026.02.01.26345288

**Authors:** Megan A. Greischar, Lauren M. Childs

## Abstract

Classic theory posits that pathogenic organisms are subject to a tradeoff between the rate and duration of transmission imposed by within-host ecology, an assumption that underpins much evolutionary theory. Here we test for a transmission-duration tradeoff using historical malaria infection data from an era prior to widespread use of antibiotics when humans were deliberately infected with malaria parasites as treatment for neurosyphilis (malariatherapy). Time series follow individual human infections until recovery or treatment with antimalarial drugs due to acute need (a proxy for virulence) and include data on the abundance of specialized transmission stages that govern parasite fitness. We fit a model to estimate initial parasite multiplication rates (PMRs) and find that initial PMRs exhibit strain-specific differences required for evolution by natural selection. However, faster PMRs extend time until recovery and enhance parasite fitness without a consistent cost of increased virulence, challenging the generality of a transmission-duration tradeoff.

## Introduction

Pathogenic microorganisms exploit resources within their hosts to multiply and transmit to new hosts. That exploitation necessarily harms hosts but is required for onward transmission, so classic theory posits that pathogenic organisms are subject to a tradeoff between the rate and duration of transmission from hosts (Frank, 1996). Under a transmission-duration tradeoff, traits that enable more efficient exploitation of hosts simultaneously render hosts more infectious but hasten the end of infections either through immune clearance leading to host recovery (Alizon, 2008) or through virulence, i.e., disease-induced mortality (Anderson & May, 1982). To balance the costs and benefits, selection is predicted to favor intermediate—rather than maximal—levels of host exploitation (Frank, 1996). Since that logic places bounds on the fitness of pathogenic organisms, the assumption of such a tradeoff enables prediction of evolutionary responses to human interventions like widespread vaccination (Gandon *et al*., 2001; Miller & Metcalf, 2022). Yet many human infections induce mortality rates lower than thought necessary to constrain pathogen evolution (Bull & Antia, 2022; Kennedy, 2023), and interactions with immunity can have complex impacts on infection duration (Bull & Lauring, 2014). Thus, the generality of a transmission-duration tradeoff remains uncertain.

The uncertainties are prominent in human malaria infections, where evidence is mixed on the relevance of a transmission-duration tradeoff to parasite evolution. Virulence scales positively with parasite biomass within the host, at least beyond a threshold parasite abundance (Mackinnon & Read, 2004; Cunnington *et al*., 2013). Heritable variation in virulence across parasite genotypes is evident from experimental rodent malaria infections (Mackinnon & Read, 1999), where variation across hosts is minimal. Parasite differences could be less important in infections of humans, who exhibit substantial variation capable of driving differences in virulence (e.g., the presence of chronic conditions like diabetes, Ch’ng *et al*. 2021, counterproductive immune responses, reviewed in Cunnington *et al*. 2013). Even with consistent differences across parasites, the risk of host mortality may be too low to drive parasite evolution (Greischar *et al*., 2016c; Kennedy, 2023). Complicating matters further, greater parasite biomass appears to slow rather than hasten the time until host recovery (Mackinnon & Read, 2004), a pattern that can emerge from interactions between parasites and immune defenses (Klein *et al*., 2014; Patterson *et al*., 2026).

Identifying the constraints on parasite evolution requires disentangling the relative importance of virulence and recovery and linking those outcomes to parasite multiplication rates (PMRs) and infectiousness. Despite a wealth of data, quantifying PMRs has proven remarkably difficult due to a recently identified methodological challenge (Greischar & Childs, 2023, 2025). Existing methods to estimate PMRs do not account for nearly periodic changes in detectability over the parasite life cycle within the blood (Fig. 1A). As parasites mature within infected red blood cells (iRBCs), those iRBCs tend to adhere to blood vessel walls (sequester) where they cannot be readily sampled, so that infection time series represent an incomplete picture of within-host ecology that limits capacity to link PMRs to the risk of mortality (Gnangnon *et al*., 2021). Parasite development within iRBCs is often synchronized across the population of parasites within an infection (Greischar *et al*., 2019b). Combined with sequestration, synchrony generates apparent fluctuations in iRBC abundance (corresponding to the 48 hour multiplicative cycle in *Plasmodium falciparum* infections, e.g., Fig. 1B), which become damped over the course of infection. As synchrony decays, the resulting increase in detectability can generate orders-of-magnitude overestimates of PMRs when using the standard approach of fitting a regression line to the log-linear expansion phase of infection (Greischar & Childs, 2023). Though these difficulties are expected to arise in diverse organisms—which often vary in their detectability throughout the life cycle and exhibit changing synchrony—the biology of *P. falciparum* gives rise to extreme bias in PMRs estimates (Greischar & Childs, 2025).

**Figure 1:**
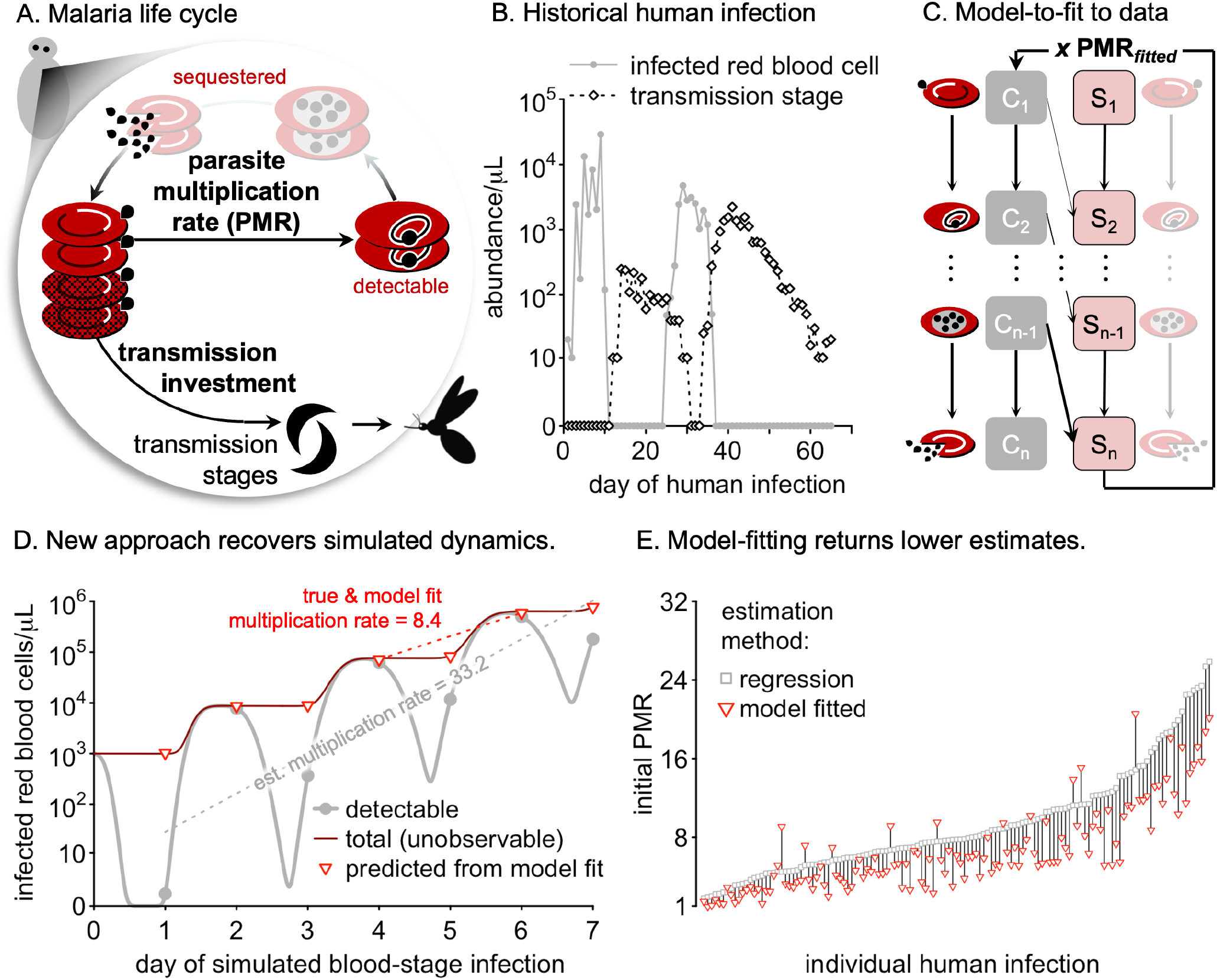
(A) Schematic representation of the parasite life cycle in the red blood cell. Within infected red blood cells, malaria parasites produce forms for either multiplication or transmission. Parasite development can be synchronized (as shown), and sampling becomes more difficult late in development when infected red blood cells sequester (shown as faded red blood cells). (B) Data from a single historical human infection include the abundance of infected red blood cells and transmission stages (patient data represented in Fig. 1C in Eichner *et al*. (2001)). (C) The model-to-fit includes two parallel tracks of parasite development within circulating (*C*, detectable) and sequestered (*S*, hidden from sampling) infected red blood cells, incorporating empirically-derived knowledge regarding the timing of sequestration. (D) Applied to simulated time series from our data-simulating model (see ‘Model fitting details’ in the Supplement), our approach is able to recover underlying dynamics infeasible with current regression methods. (E) When fit to human infection data, our novel method typically returns lower initial multiplication rates than the regression method.

Overcoming these challenges in *P. falciparum* infections is an opportunity to uncover the links between multiplication rates and the degree and duration of host infectiousness. Such analysis is possible due to the existence of historical human data collected prior to widespread use of antibiotics, when neurosyphilis infections were treated by deliberate infection with human malaria parasites (including *P. falciparum*, Collins & Jeffery 1999). Since ‘malariatherapy’ time series— near daily abundance of iRBCs and transmission stages (Fig. 1B)—were collected in the course of treatment, they are deemed ethical for retrospective analysis (Weijer, 1999; Humphreys, 2003). There is no evidence that coinfection with neurosyphilis altered the early dynamics of malaria infections (e.g., via immune priming) since malariatherapy infections follow similar trajectories compared to modern data from controlled human malaria infection trials during the initial phase of infection (Greischar & Childs, 2023). While this initial phase is all that can be followed in modern trials, malariatherapy infections cover the full duration of infections. Antimalarial drugs were used sparingly on these infections, since the combination of parasite multiplication and fevers produced clinical improvements in neurosyphilis (Collins & Jeffery, 1999; Humphreys, 2003). Many infections were untreated, providing data and time until host recovery, and infections that were drug treated due to acute need provide a proxy for virulence, since hosts would presumably have died from their malaria infections had drugs not been administered, putting an end to transmission opportunities. The fitness implications can be estimated using a an empirically-derived curve relating *P. falciparum* transmission stage abundance to the probability of transmission to mosquitoes (Huijben *et al*., 2010).

To test for a transmission-duration tradeoff in human malaria parasites, we validate and employ a novel model-fitting approach to recover unbiased PMRs from malariatherapy infections. We focus on initial PMRs since early infection dynamics are thought to have a disproportionate impact on transmission success (Greischar *et al*., 2016a) and infection duration (Klein *et al*., 2014), and because initial PMRs reflect maximal rates of multiplication before dwindling red blood cells and upregulation of immunity curtail parasite population growth. Our analysis reveals significant and consistent differences in PMRs across parasite strains despite considerable variability across hosts, suggesting potential for heritable parasite variation. We find three lines of evidence against a classic transmission-duration tradeoff: (1) initial multiplication rates had no detectable impact on virulence, whether quantified as the proportion of infections that were acutely treated or as the time until acute treatment; (2) for acutely treated infections (a proxy of virulence), faster initial within-host multiplication corresponds with reduced—rather than increased—probability of transmission; and (3) for untreated infections, faster multiplication is associated with longer times until host recovery, a short-term increase in the average probability of transmission, and a significant net increase in the potential for transmission over the course of infection. Moreover, in our dataset, we find that hospital has a significant impact on the odds of acute treatment—overwhelming any signal of differences across parasite strains—while strain-specific differences in recovery remain consistent. Thus, our analysis suggests that greater, rather than intermediate, rates of within-host multiplication extend infection duration and enhance parasite fitness.

## Methods

To account for *P. falciparum* biology that causes overestimated PMRs by standard regression methods (Greischar & Childs, 2023), we fit an ordinary differential equation model (Fig. 1C) to simulated time series. Briefly, the model-to-fit (Archer *et al*., 2018; Greischar & Childs, 2023, 2025) tracks parasite development through two parallel tracks—circulating versus sequestered iRBCs— assuming that iRBCs sequester as they mature according to an empirically-derived relationship for *P. falciparum in vitro* (Kriek *et al*., 2003). We first validate our model-fitting approach against simulated time series from a different, mechanistic model that recapitulates the challenges of observed data, i.e., sequestration of iRBCs later in development and decay of synchrony (Fig. 1D). We then apply the model-fitting method to malariatherapy data to investigate the links between initial PMRs, virulence, recovery, and parasite fitness. We confirm the robustness of our results with additional comparisons, including analyzing virulence and recovery as competing risks (see ‘Additional supporting analyses’ in the Supplement).

### Malariatherapy strains

The dataset consists of 305 individual *P. falciparum* infections from three strains: El Limon (79 infections), McLendon (141 infections), and Santee Cooper (85 infections), along with additional 17 infections with strains that were used rarely (none represented in more than 5 infections) and not considered in the present study. Of the three well-represented strains, two were isolated in South Carolina (McLendon and Santee Cooper) in 1940 and 1946, respectively, while the third strain (El Limon) was isolated in Panama in 1948 (reviewed in Collins & Jeffery 1999). Malaria was endemic to the US during this time (Andrews *et al*., 1950), though in decline due to widespread eradication efforts led by what is now the Centers for Disease Control and Prevention (CDC), which was originally established to combat malaria transmission in the US (CDC, 2026). Although methods were not yet available to genetically sequence isolates at the time, a malariatherapy strain almost certainly does not represent a single genotype. Upon the advent of genetic sequencing, it was shown that parasite strains isolated from human infections often contain multiple distinct parasite genotypes (reviewed in McKenzie *et al*., 2008). However, genetic differences among strains are thought to become eroded within a locale due to frequent co-transmission to mosquitoes and concomitant interbreeding of strains, while differences are expected to be maintained across larger geographic distances (McKenzie *et al*., 2008). As a result, the genetic variation within a strain is smaller than the variation across strains isolated in different years or from distinct geographic regions (“effective clonality”, Mu *et al*., 2005). The variability within a malariatherapy strain is similar to the within-strain variation exhibited by putatively ‘clonal’ strains artificially cultivated with modern methods, where genetic variation accumulates rapidly even with tightly controlled artificial culturing methods (Freitas-Junior *et al*., 2000; Liu *et al*., 2025; Brown *et al*., 2026). Given the prevalence of genetic variation within a strain, the key determinant of potential for evolutionary change is whether there are consistent differences across parasite strains, distinct from variability across hosts.

### Defining virulence, recovery, and censored data

Estimating the duration of infection necessitates considering two competing hazards by which infections could end, virulence versus recovery. Early reports found that malaria-induced mortality occurred in 1% to 9% of malariatherapy infections, with lower case fatality rates achieved by screening patients for risk of complications (reviewed in Austin *et al*., 1992) and by application of antimalarial drugs to prevent adverse health outcomes (de Montjoie Rudolf, 1927; Weiss, 1940; Collins & Jeffery, 1999; Humphreys, 2003). Consistent with those practices, none of the malariatherapy infections in the dataset we utilize here ended in patient death. To assess the risk of infections ending through virulence, we assume malaria-induced mortality would have occurred if patients had not received antimalarial drugs for acute need (henceforth “acutely treated infections”). To isolate cases of acute need, we selected infections that were treated when circulating iRBC abundance ≥ 1000*/µL*, a threshold recommended for treatment of symptomatic infections in modern times (Färnert *et al*., 1997).

Distinct from these cases, many malariatherapy infections were allowed to run their course without drug treatment, providing data with which to estimate the time until host recovery. We calculated the time until host recovery as the last day on which an iRBC abundance above the detection threshold was reported for patients who were not drug treated or subject to secondary inoculation with another *P. falciparum* strain, a practice used for patients still not cured of their syphilis infections (McKenzie *et al*., 2008). We refer to infections that were not treated with drugs or secondary inoculation as “untreated”. Note that for clarity throughout, iRBC abundance excludes transmission stages.

We classify other infections as “censored”, since patients were subject to additional interventions so that the time until last detectable iRBCs cannot be estimated with confidence. This censoring of recovery times occurred via secondary inoculation or drug treatment not due to acute need (circulating iRBC abundance *<* 1000*/µL*), e.g., after the last day of detectable iRBC abundance, presumably to prevent recrudescence.

### Competing risk analysis

To ensure that our results are robust to potential bias in data selection, censoring, and choice of virulence proxy, we perform non-parametric comparisons of competing risks (i.e., recovery versus virulence) using cumulative incidence functions following the methods in (Zhang, 2017). This type of analysis enables us to use information from censored infections (i.e., those subject to secondary inoculation or non-acute drug treatment) and to account for the fact that one outcome of interest (virulence) precludes the other (recovery), providing more robust estimates of risks (Basak *et al*., 2021). We only analyze differences when all groups being compared each contain at least one infection classified as ending in recovery and at least one infection classified as ending in virulence.

### Model-fitting approach

To estimate initial PMRs, we fit an ordinary differential equation model that describes parasite development within red blood cells for *P. falciparum* along two parallel tracks, for circulating and sequestered iRBCs, respectively (Archer *et al*., 2018; Greischar & Childs, 2023, 2025). The fitted ODE model—distinct from the data-simulating model—uses 5 parameters to describe dynamics of circulating and sequestered iRBCs: the population growth rate, PMR_*fitted*_ (assumed to be constant, since we focus on the initial log-linear expansion phase of infection); the number of developmental age compartments, *n* (where lower *n* enable more rapid decay of synchrony through time); and 3 parameters to specify the initial iRBC population—the total initial iRBC abundance, *T* ; the median *ω*_*med*_ and the width *ω*_*range*_ of a symmetric Beta distribution to form the initial age distribution, which specifies the level of synchrony (Greischar *et al*., 2024). Synchrony decays more slowly as the number of developmental age compartments *n* increases, since a shrinking fraction of the population traverses the *n* developmental age compartments considerably faster or slower than the mean rate *ε* = *n/*48, where 48 hours is the time from red blood cell invasion to bursting in *P. falciparum* (reviewed in Mideo *et al*., 2013). The model-to-fit is given by:

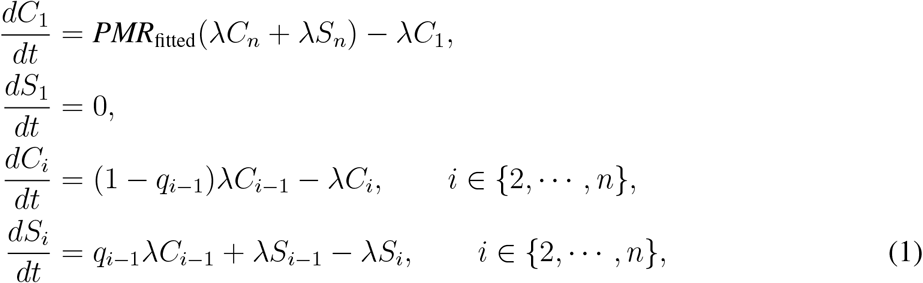

where *C*_*i*_ represents the *i*th circulating age compartment and *S*_*i*_ the *i*th sequestered age compartment. The fraction of maturing circulating iRBCs that enter the sequestered track of development (*q*_*i*_) is given by

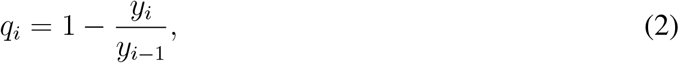

where *y*_*i*_ = 1 − *g*_*i*_ such that *g*_*i*_, the fraction of parasites assumed to be sequestered by time *t*_*i*_, is given by

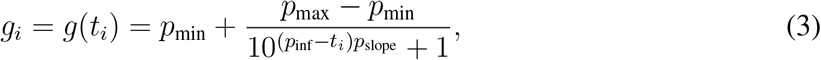

where *p*_min_ is the minimum level of sequestration, *p*_max_ is the maximum, *p*_inf_ determines the inflection point, *p*_slope_ determines the slope at the inflection point, and *t*_*i*_ is the time during intraerthyocytic development of the beginning of age compartment *i*. We assume the values fitted by Archer *et al*. (2018) to *P. falciparum* data from artificial culture (Kriek *et al*., 2003), such that *p*_min_ = 11.3869*/*467.6209, *p*_max_ = 1, *p*_inf_ = 18.5802, and *p*_slope_ = 0.2242. We used optimization with the Nelder-Mead algorithm to minimize the sum of squared errors (SSE) of the log_10_ model predicted and observed (or generated in the case of simulated time series) abundance of circulating iRBCs, adding one to the abundance before logging to ensure no zeros (see ‘Model fitting details’ in the Supplement).

### Validation of approach

#### Simulated time series & data-simulating model

To recapitulate the challenges of observed data, we modified a detailed mechanistic model of malaria infections (Pak *et al*., 2024) parameterized for human *P. falciparum* infections following (Greischar *et al*., 2019a, details in Fig. S1). This model incorporates the two features we identified as generating PMR overestimates (Greischar & Childs, 2025): (1) developmental sampling bias, in that only the first *k* developmental age classes are detectable; and (2) variation in the duration of parasite development within iRBCs, allowing synchrony to decay through time (Greischar *et al*., 2019b). However, compared with the model-to-fit, synchrony will decay slightly faster in this data-simulating model due to the inclusion of a brief merozoite stage, in which merozoite lifespans follow an exponential distribution to mimic *P. falciparum* dynamics *in vitro* (Boyle *et al*., 2010).

From our data-simulating model, we simulate *P. falciparum* time series from the log-linear expansion phase of infection (the first seven days, to mirror the data, Fig. 1C). For the first week of simulated time series, we calculate the total abundance of iRBCs (detectable and sequestered) as *I*(*t*) = *I*_1_(*t*) + *I*_2_(*t*) + … + *I*_*n*_(*t*), where subscripts index the developmental age class of iRBCs, representing time since parasite invasion (see ‘Model fitting details’ in the Supplement). We compared the true PMR (i.e., the PMR obtained with perfect information, calculated as *I*(*t* + 2)*/I*(*t*) for day *t* since the duration of the multiplicative cycle is 2 days), against PMR estimates obtained from simulated time series of detectable iRBCs using either regression or model-fitting.

### Fitting to malariatherapy data

Of the full dataset consisting of one of the three well-represented strains (305 infections), we fit 161 infections with the model-to-fit and 131 infections are used in our main analysis. The 161 infections fit represent all of the time series meeting the following three criteria: (1) the infection was not treated with drugs during the first seven days; (2) there were no missing values during the first seven days of data; (3) the peak of infection occurred no earlier than day 6, where the peak of infection was defined following Eichner *et al*. (2001) as the first local maximum in iRBC abundance that was preceded and followed by 6 lower values. Early drug treatment or an early peak in infection would violate the assumption of log-linear (i.e., constant) population growth, an assumption of previous methods to estimate PMR (e.g., regression, Simpson *et al*., 2002), so we follow previous studies and exclude these infections from our analysis. Thus, the full dataset included 144 infections that violated one or more of criteria (1)-(3) with reasons shown in Fig. S2.

Of these 161 fitted infections, we first excluded 4 which exhibited consecutive zeros in their iRBC time series, a situation that falls outside the scenarios our model-to-fit can parsimoniously describe, leaving 157 infections. Based on poor fit to data, we also excluded an additional 26 fits with SSE ≥ 1 (Fig. S3). See ‘Additional supporting analyses’ in the Supplement for details and demonstration of qualitatively identical patterns with a higher cutoff.

### Estimating parasite fitness

#### Infectiousness

To estimate host infectiousness, we calculate the probability of infecting mosquitoes at each time point for which transmission stages were censused using the curve empirically-derived from *P. falciparum* infections by Huijben *et al*. (2010):

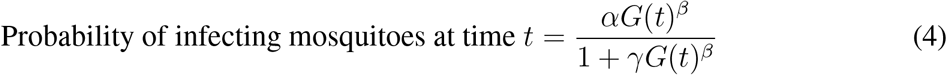

where *G* represents abundance of transmission stages per microliter, *α* = 0.03, *β* = 0.6, and *γ* = *α/*0.85. To account for missing values, we calculate the average probability of transmission for patients that either recovered or were acutely drug treated, excluding any days on which transmission stage abundance was not reported. All infections included in our analysis had at least one non-missing count of transmission stage abundance. For acutely treated infections, we calculated average probability of transmission through the first day of drug treatment. For patients that recovered without intervention, we calculate the average probability of transmission over the entire duration of infection.

#### Transmission potential

Transmission potential represents parasite fitness analogous to the metric used in HIV infections (Fraser *et al*., 2014), i.e., the average probability of infecting mosquitoes multiplied by the duration of infectiousness, calculated from the first day of blood stage infection to either first day of drug treatment (for acutely treated infections) or to last detection of transmission stages (for untreated infections). Infections consisting of only zero counts of transmission stage abundance were included in analyses and assigned a transmission potential of zero.

## Results

### Validation of model-fitting approach

Our model fitting approach recovers accurate PMRs from simulated time series (Fig. 1D). In contrast, standard regression methods returned nearly a four-fold higher multiplication rate than the true value for simulated time series. Among the 131 infections that meet our criteria (see Methods for details), we find that our approach typically returns lower values than standard regression methods (Fig. 1E), consistent with prior findings that regression is prone to overestimation (Greischar & Childs, 2023).

### Variation within and across parasite strains

The human malaria infections we fit exhibited considerable variation in their dynamics, including the time of peak infection (Fig. 2, Fig. S4). We find the McLendon strain exhibits slower initial PMRs and peak iRBC abundance, while El Limon and Santee Cooper exhibit similar median values (Fig. 2C, D). There are no apparent differences in the timing of acute drug treatment across strains (Fig. 2E), but untreated McLendon infections are shorter (Fig. 2F) and exhibit reduced transmission potential (Fig. 2G). With respect to time until recovery, we find that faster PMRs are associated with longer infections—the opposite pattern to a transmission-duration tradeoff, under which we would expect that faster within-host multiplication would reduce infection duration. In subsequent comparisons, we conclude differences are significant if the mean of one group falls outside the bootstrapped 95% confidence interval of another group.

**Figure 2:**
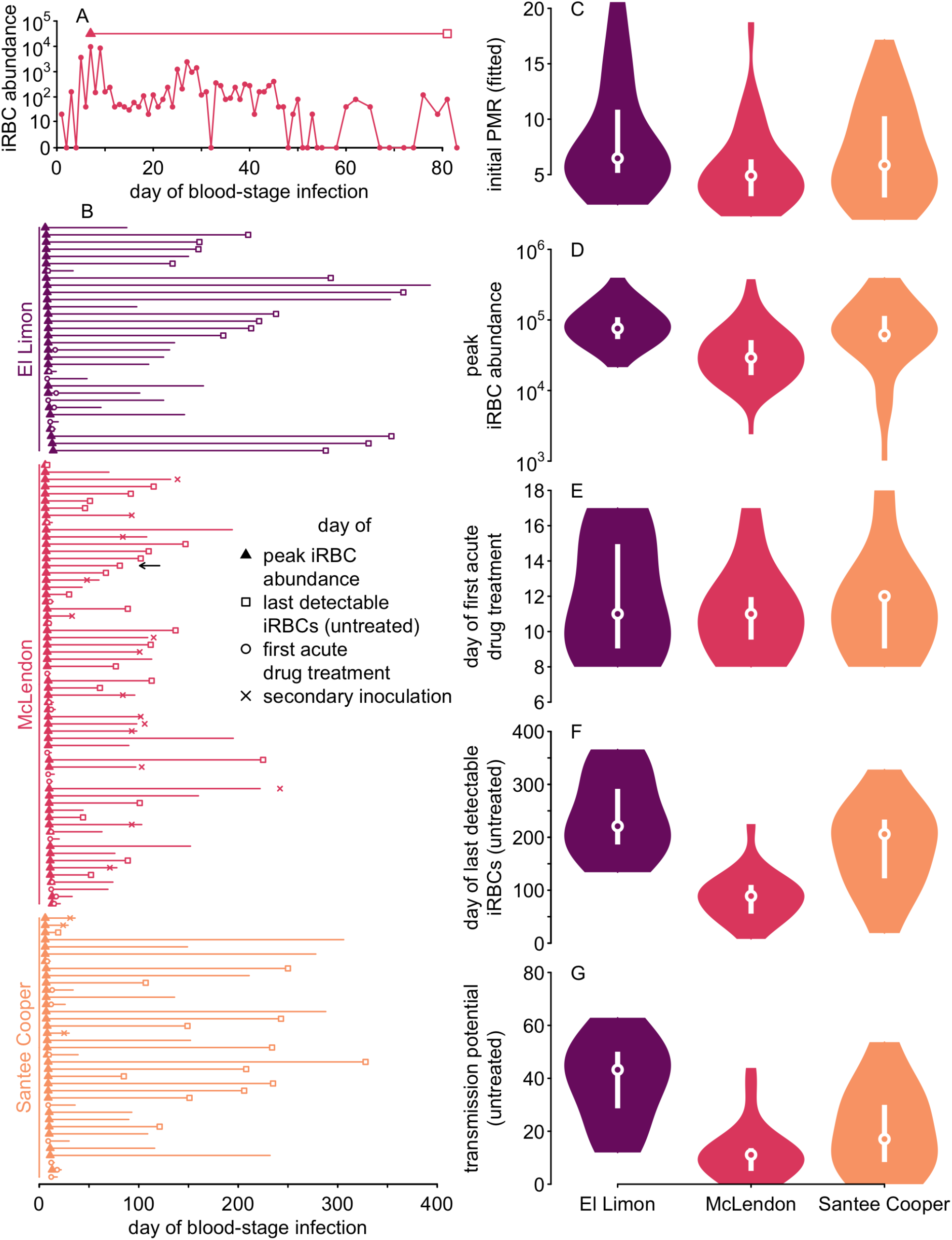
The three *P. falciparum* strains well-represented in the data exhibit substantial variation in their dynamics. (A) Infection time series encompass nearly daily samples, with a closed triangle to indicate the peak of infection. The end of the horizontal line indicates the last day on which iRBCs were detected, with an open square to indicate that this infection was never drug-treated. (B) The time from peak to end of infection is shown with horizontal line segments grouped for each of the three common strains, ordered from earliest to latest peak. The time of peak iRBC abundance is only indicated with a closed triangle if it occurred prior to drug treatment (first drug treatment shown with an open circle), while ‘x’ denotes the day on which another *P. falciparum* strain was inoculated. Violin plots compare values across the three strains, including (C) initial fitted PMRs (for all infections meeting the inclusion criteria defined in main text), (D) peak iRBC abundance*/µ*L (for all infections untreated on or before peak infection), (E) the day of first acute drug treatment (for acutely treated infections only), (F) the day of last detectable iRBCs (for untreated infections only), and (G) the transmission potential (also for untreated infections only, see Methods for details). For each violin, an open point indicates the median while white bars denote the first and third quartile.

### Faster PMRs extend time until recovery with no impact on virulence

We find that infections with faster initial PMR values exhibit larger peak iRBC abundance, among infections that were not drug treated at or before their peaks (Fig. 3A). Infections with the McLendon strain exhibit both slower initial PMRs and reduced peak abundance. Since we cannot rule out interactions between strain and hospital (see ‘Additional supporting analyses’ in the Supplement), we compute means and 95% confidence intervals separately for each strain-hospital combination that included at least three infections. We find that the positive correlation between initial PMR and peak iRBC abundance is not solely driven by differences among strains, because the pattern remains significant in the studentized residuals (the residual of a value with respect to the mean for the strain-hospital combination, divided by the standard deviation for that combination, Fig. 3B). Thus, infections that have faster initial PMRs than are typical for that strain and hospital also have atypically greater peak iRBC abundance (Fig. 3A-B).

**Figure 3:**
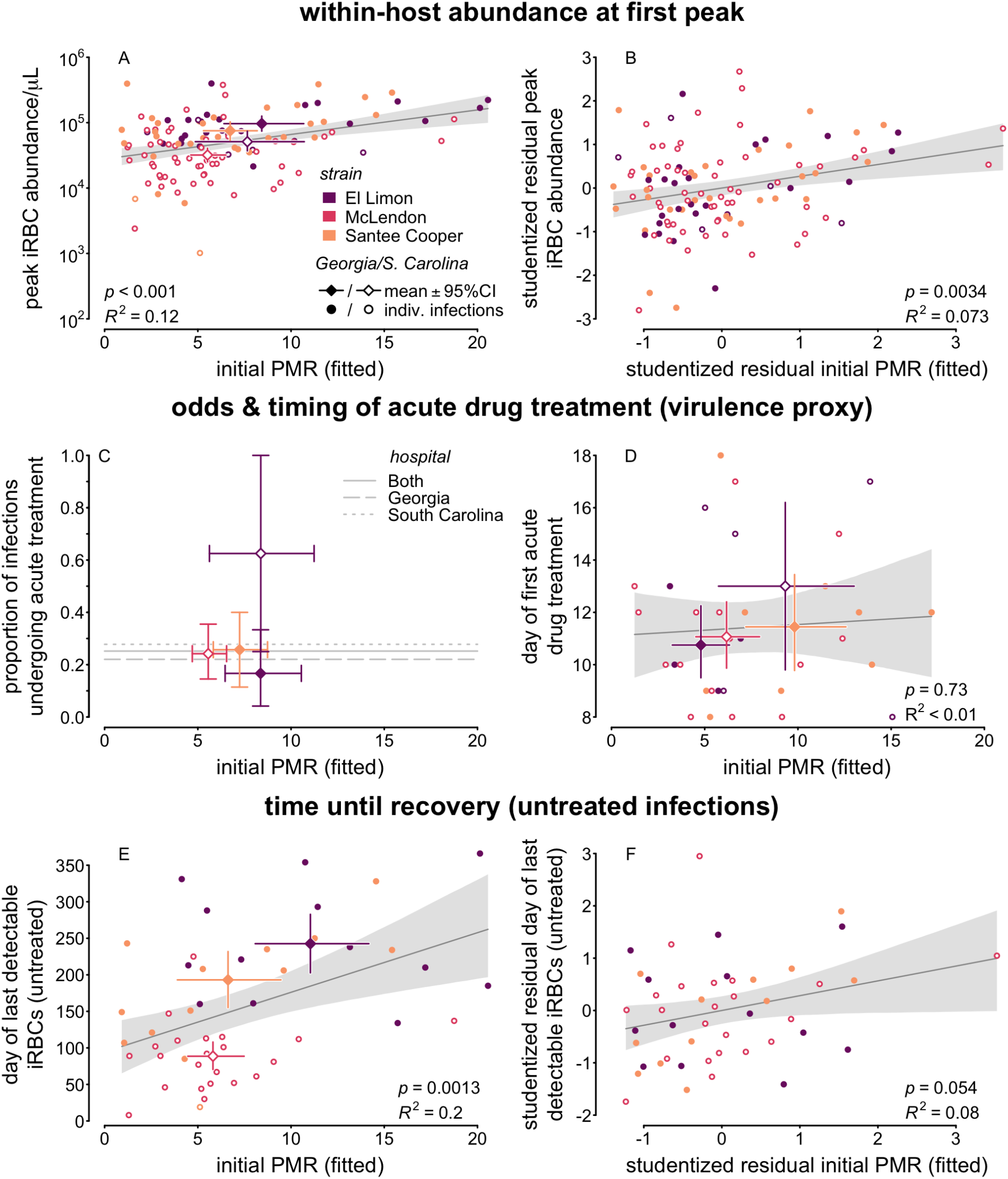
Faster initial PMRs correlate with higher peak abundance and longer times until host recovery, but not odds or timing of acute drug treatment. Linear regressions are shown as dark gray lines, with 95% confidence bounds shown as a lighter gray polygon. Individual closed points indicate values from individual infections from Georgia State Hospital while open points refer to infections from South Carolina State Hospital, with diamonds to indicate strain/hospital means*±*95% confidence intervals (only calculated for combinations consisting of more than 2 infections). (A) Individual infections exhibit a significant positive relationship between initial PMR and peak iRBC abundance (on a log_10_ scale). (B) A significant positive relationship also appears in the studentized residuals. (C) Despite some differences across strains and hospital, the initial PMR has no discernible relationship with the proportion of infections that were acutely drug treated, (D) nor the time until acute drug treatment. (E) The time until host recovery among untreated infections exhibits a positive correlation with initial PMR, (F) and that pattern is reflected among the studentized residuals.

We find no detectable relationship between initial PMRs and either the proportion of infections undergoing acute treatment (Fig. 3C) or the time until acute treatment (Fig. 3D). El Limon infections at South Carolina State Hospital were significantly more likely to be acutely treated than El Limon infections at Georgia State Hospital (see ‘Additional supporting analyses’ in the Supplement). Despite significant differences in PMRs across strains (and across hospitals), we find no significant differences in the timing of acute treatment (overlapping confidence intervals in Fig. 3D). However, we find a strongly significant positive correlation between initial PMR and time until recovery among untreated infections, with strains deviating from the average along similar lines to peak abundance: McLendon infections tend to multiply more slowly and are shorter than average across all included untreated infections, while El Limon infections multiply faster than average with longer times until recovery (Fig. 3E).

Though untreated infections can last hundreds of days, our analysis suggests that PMRs during the first week of infection can explain nearly 20% of the variation in infection length (*R*_2_ = 0.2, Fig. 3E). To determine whether the correlation between initial PMRs and time until recovery could be driven by strain-specific differences, we again investigate patterns in the studentized residuals, finding that infections with faster initial PMRs than are typical for their strain and hospital, also last longer than is typical (Fig. 3F). Thus our results do not support a key assumption of transmission-duration tradeoffs (whether mediated through virulence or recovery), that faster within-host multiplication and higher abundance abbreviates infections. Rather, we find that faster PMRs either have no impact (on virulence) or extend the time until clearance. Taken together, our findings suggest that by multiplying faster and achieving greater peak abundance within the host, malaria parasites prolong infection duration.

### Competing risks of virulence & recovery

We find consistent strain-specific differences in the risk of recovery but not virulence, suggesting that environmental factors such as hospital modulate virulence. To maximize sample sizes, we first performed additional comparisons using the full dataset consisting of the three well-represented strains (Table S1). Separating the full dataset by hospital, we find consistent strain-specific differences in recovery but not virulence (Fig. S5A, B). Focusing on the El Limon strain that was well-represented at both hospitals, we find significant differences in virulence but not recovery by hospital (Table S1). We then re-ran comparisons with our fitted dataset where sample sizes permitted (Table S2). In the fitted data, we find highly significant strain-specific differences in the risk of recovery with McLendon strain infections experiencing higher risk of recovery than the other two strains (Fig. S5C).

### Faster initial PMRs enhance parasite fitness in untreated infections

To investigate the links between within-host multiplication and parasite fitness, we examine the dynamics of production of the transmission stages required for onward transmission to mosquitoes. We find a correlation, albeit nonsignificant, between faster initial PMRs and delayed detection of transmission stages in acutely treated infections (Fig. 4A), for infections in which transmission stages were detectable at or prior to drug treatment. No such pattern exists for untreated infections (Fig. 4B), raising the possibility that risk to parasites of acute drug treatment (and virulence) could be mediated by investment into transmission stage production rather than directly through PMRs.

**Figure 4:**
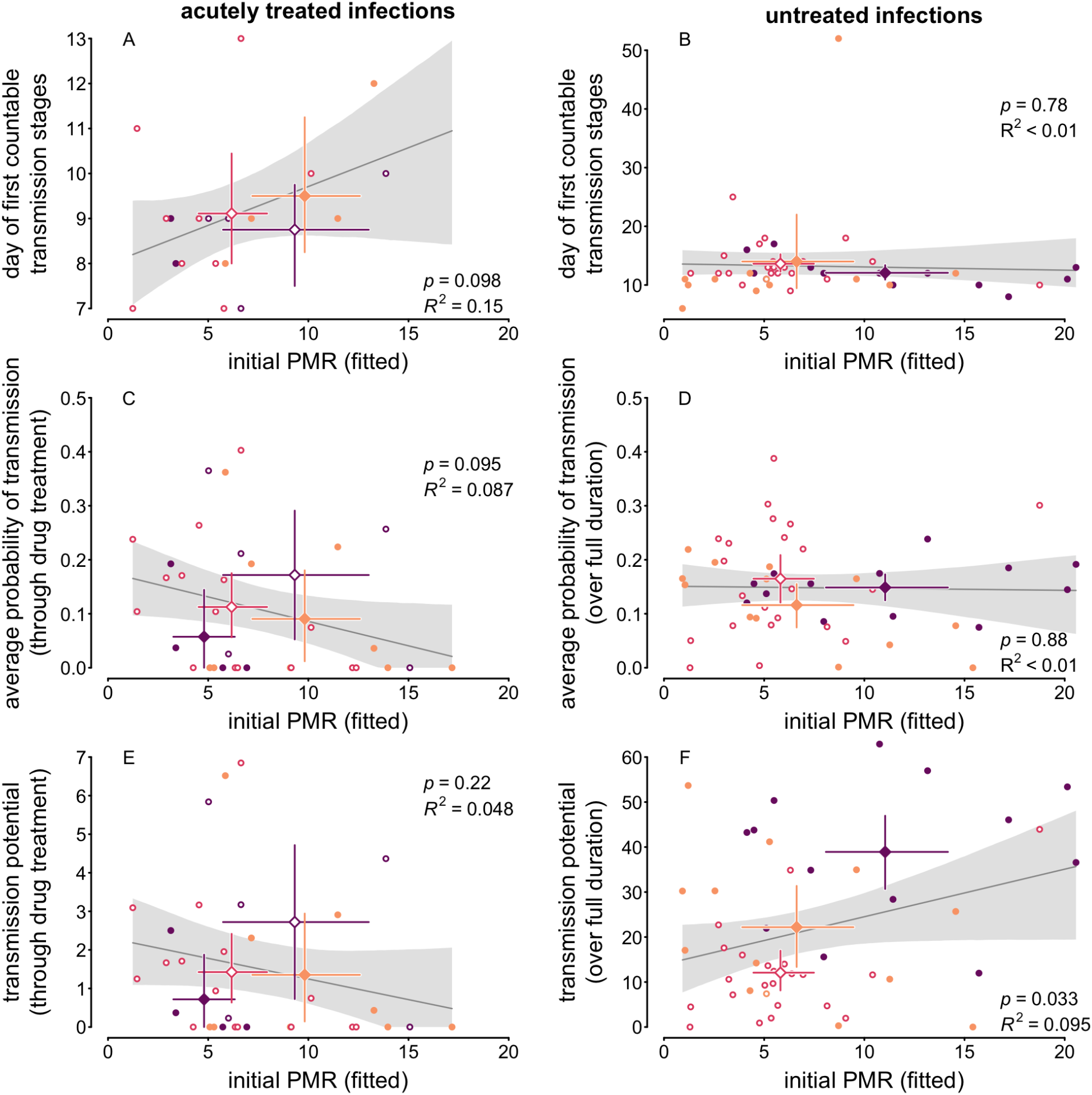
Faster initial PMRs reduce onward transmission in acutely treated infections while enhancing transmission potential in untreated infections. Linear regressions are shown as dark gray lines, with 95% confidence bounds shown as a lighter gray polygon. (A) Faster PMRs are associated with longer delays in transmission stage production in acutely treated infections, while (B) the timing of early transmission stage production remains similar as PMR increases in untreated infections. (C) Accordingly, the probability of transmission (averaged through the day of acute drug treatment) declines with initial PMR in acutely treated infections. (D) In untreated infections, initial PMRs show no consistent association with the probability of transmission averaged over the full duration of the infection. (E) For acutely treated infections, the transmission potential through drug treatment shows no correlation with initial PMRs. Note that bootstrapping linear regressions can result in slightly negative values of transmission potential. Since those are biologically impossible and an artefact of using linear regression rather than a more complicated model, we show only the portions of the confidence interval at or above zero. (F) In contrast, the transmission potential for the full duration of untreated infections increases with initial PMR. Note that full duration here refers to the timespan over which transmission stage counts were reported, which could have differed from the time until day of last detectable iRBCs.

For acutely treated infections, faster initial PMRs were weakly associated with delays in transmission stage production, nonsignificantly reducing the average probability of transmission (Fig. 4C). In contrast, the probability of transmission averaged over the full duration of untreated infections shows no correlation with initial PMRs (Fig. 4D). In line with reported timing for transmission stage production and longevity Lensen *et al*. (1999); Eichner *et al*. (2001); Smalley & Sinden (1977), our analysis suggests that rapid initial multiplication supports subsequent increased transmission stage production which may persist for weeks (Fig. S6). After that period, the signature of initial PMR decays quickly, and the average probability of transmission is dominated by later fluctuations in abundance of iRBCs (e.g., Fig. 2A) and the transmission stages they produce. For acutely treated infections, the median time until first drug treatment was 11 days, so that—if drugs had not been administered—host mortality would be expected to terminate infections before any transmission gains from faster within-host multiplication could be realized.

To enable comparison with evidence for transmission-duration tradeoffs in HIV (Fraser *et al*., 2014), we next calculate the transmission potential. In acutely treated infections, faster initial PMRs are associated with a slight reduction in transmission potential (Fig. 4E), corresponding to the delay in transmission stage production and subsequently reduced probability of transmission. In untreated infections, the transmission potential increases significantly with initial PMR (Fig. 4F). Therefore, our results are consistent with the idea that faster PMRs increase parasite fitness in untreated infections by extending the duration of host infectiousness.

The patterns we find for fitness in *P. falciparum* infections also diverge from those expected under a transmission-duration tradeoff. The expected patterns are exemplified in HIV infections, where transmission potential is maximized at intermediate viral loads (Fraser *et al*., 2014). We rule out such a unimodal relationship by including a quadratic term in the regression of transmission potential against PMR, finding no significant improvement to fit compared with our linear regression (Fig. S7), as would be expected under a transmission-duration tradeoff. No significant association emerges between PMRs and transmission potential for acutely treated infections. In untreated infections, transmission potential increases with initial PMR, rather than attaining an optimum at intermediate rates of within-host multiplication. It is unlikely that a decline in transmission potential would emerge at still faster PMRs, since the initial PMRs we estimate from the malariatherapy data already span the full plausible range given current understanding of *P. falciparum* biology. Artificial culture experiments show that the median number of daughter parasites produced by a bursting iRBC ranges from approximately 16-18 (Reilly *et al*., 2007), and maximal PMRs would be expected to fall below median burst sizes since not every daughter parasite will successfully invade an RBC. Thus, our results argue against the existence of an intermediate optimum for PMRs.

## Discussion

Though the assumption of a transmission-duration tradeoff represents a mainstay of models aimed at predicting the evolution of pathogenic organisms, we find that such a tradeoff is not general. Under a transmission-duration tradeoff like those reported for HIV (Fraser *et al*., 2014) and dengue (Ben-Shachar & Koelle, 2018), traits that enhance within-host abundance should increase instantaneous rates of transmission while shortening infection duration. In contrast to evidence from HIV and dengue, we find no evidence that intermediate PMRs maximize parasite fitness, but rather show that parasite transmission potential is higher either at slow initial PMRs (for acutely treated infections) or fast initial PMRs (for untreated infections). The marked lack of evidence for a transmission-duration tradeoff in our analysis raises the possibility that by producing specialized transmission stages, malaria parasites may be able to sidestep the constraints that limit other pathogenic organisms, like viruses.

Our analysis supports the idea that virulence can be costly, since much greater transmission potential is possible from untreated versus acutely treated infections. However, we find no evidence that faster PMRs hasten virulence and no consistent differences in virulence across parasite strains. Notably, the risk of mortality due to malariatherapy was regarded at the time as a property of patient rather than parasite characteristics (de Montjoie Rudolf 1927, reviewed in Austin *et al*. 1992). Consistent with that view, we find that strain differences in virulence depend on environmental context, here, the hospital where malariatherapy infections were administered. The available data do not reveal the causes of variation across hospitals, but two possibilities are differences in medical practices—e.g., how patients were assessed for adverse events—or variation in the demographic characteristics of populations served by the two hospitals. Notably, South Carolina State Hospital reported elevated rates of mortality from malariatherapy infections compared to the national average (Pelletier, 2025). The dataset used in our analysis lacks information on any patient demographic characteristics, so we cannot test how those characteristics impact virulence, but we find elevated rates of acute drug treatment (our proxy for virulence) at South Carolina State Hospital, even controlling for parasite strain. The importance of environmental context in driving patterns of virulence is in line with evidence from other systems, including SARS-CoV-2 where both properties of human populations (such as age structure) and healthcare infrastructure are known to have an outsize impact on the risk of disease-induced mortality (Alizon & Sofonea, 2021). Accordingly, our analysis of *P. falciparum* infections uncovered no consistent strain-specific differences in virulence that would be necessary for virulence to evolve. In contrast, we found that strain-specific differences in the risk of recovery were consistent despite variation across patients and hospitals, with faster-multiplying strains capable of persisting longer within the host. This pattern also held within strains, such that faster-than-average PMRs correlated with longer-than-average infections. These results align with both model predictions (Klein *et al*., 2014; Patterson *et al*., 2026) and field data suggesting that greater parasite biomass prolongs immune clearance (Mackinnon & Read, 2004). Altogether, our findings suggest that faster within-host multiplication carries an inconsistent cost while achieving a consistent and significant benefit by prolonging infection duration.

Yet the assumption of a transmission-duration tradeoff remains nearly ubiquitous in general models of pathogenic organisms (reviewed in Bull & Lauring 2014; Bull & Antia 2022). Notable examples include theory for evolution in response to vaccination (Gandon *et al*., 2001; Miller & Metcalf, 2022) and for the evolution of transmission prior to symptoms (Saad-Roy *et al*., 2020, 2021a,b). Both evolutionary questions are highly relevant to human malaria parasites, since two vaccines are currently approved for use against *P. falciparum* (The World Health Organization, 2025) and considerable transmission occurs from malaria infections with no obvious symptoms (Lindblade *et al*., 2013). Extending existing theory that relies on a transmission-duration tradeoff is nontrivial, since some kind of constraint on parasite fitness is needed to prevent evolution towards ever more aggressive exploitation of hosts.

Recent theory suggests that a reproduction-survival tradeoff, rather than a transmission-duration tradeoff, can constrain the evolution of human malaria parasites (Patterson *et al*., 2026). When specialized transmission stages are required for spread to new hosts, the production of those transmission stages reduces the capacity to multiply within the present host, leaving parasites vulnerable to immune clearance. Our present analysis uncovers patterns consistent with those modeling results, including that both transmission potential and the time until host recovery tend to increase with faster within-host multiplication. Testing those predictions directly would be possible by extending the methods we develop here to estimate how PMRs and total iRBC abundance (circulating and sequestered) vary over the course of infection. With reconstructed within-host dynamics, methods exist to estimate investment into transmission stage production (Eichner *et al*., 2001; Greischar *et al*., 2016b). That expanded analysis would reveal how transmission investment strategies impact transmission potential.

While heritable variation in parasite growth capacity has been demonstrated *in vitro* (Reilly, 2007), determining whether that parasite variation is expressed *in vivo* has remained out of reach due to methodological challenges surrounding PMR estimation (Greischar & Childs, 2023, 2025). Despite considerable variation in PMRs across individual patients, we find consistent, significant strain-specific differences that are likely to have consequences for parasite fitness, creating potential for evolution by natural selection. PMRs could therefore evolve in response to human efforts to disrupt malaria transmission, but not in ways that are readily predicted by existing theory. Locating the precise tradeoffs that constrain parasite evolution remains an urgent practical question.

## Data Availability

All data in the present study are available upon reasonable request to the authors and will be supplied as supplementary files upon publication.

## Acknowledgments

We thank Nicole Mideo, Sarah Reece, and members of their labs for useful discussions, as well as Lucas Nell, Weixin Du, Zhenying Chen, and Louie Gold.

## References

Alizon, S. (2008). Transmission-recovery trade-offs to study parasite evolution. The American Naturalist, 172, E113–E121.

Alizon, S. & Sofonea, M. T. (2021). SARS-CoV-2 virulence evolution: Avirulence theory, immunity and trade-offs. Journal of Evolutionary Biology, 34, 1867–1877.

Anderson, R. & May, R. (1982). Coevolution of hosts and parasites. Parasitology, 85, 411–426.

Andrews, J. M., Quinby, G. E. & Langmuir, A. D. (1950). Malaria eradication in the United States. American Journal of Public Health and the Nations Health, 40, 1405–1411.

Archer, N. M., Petersen, N., Clark, M. A., Buckee, C. O., Childs, L. M. & Duraisingh, M. T. (2018). Resistance to Plasmodium falciparum in sickle cell trait erythrocytes is driven by oxygen-dependent growth inhibition. Proceedings of the National Academy of Sciences of the United States of America, 115, 7350–7355.

Austin, S. C., Stolley, P. D. & Lasky, T. (1992). The history of malariotherapy for neurosyphilis: Modern parallels. JAMA: The Journal of the American Medical Association, 268, 516–519.

Basak, R., Mistry, H. & Chen, R. C. (2021). Understanding competing risks. International Journal of Radiation Oncology Biology Physics, 110, 636–640.

Ben-Shachar, R. & Koelle, K. (2018). Transmission-clearance trade-offs indicate that dengue virulence evolution depends on epidemiological context. Nature Communications, 9.

Boyle, M. J., Wilson, D. W., Richards, J. S., Riglar, D. T., Tetteh, K. K. A., Conway, D. J., Ralph, S. A., Baum, J. & Beeson, J. G. (2010). Isolation of viable Plasmodium falciparum merozoites to define erythrocyte invasion events and advance vaccine and drug development. Proceedings of the National Academy of Sciences of the USA, 107, 14378–14383.

Brown, N., Luniewski, A., Yu, X., Warthan, M., Liu, S., Zulawinska, J., Ahmad, S., Prasad, N., Congdon, M., Santos, W., Xiao, F. & Guler, J. L. (2026). Replication stress increases de novo CNVs across the malaria parasite genome. Nucleic Acids Research, 54, gkaf1436.

Bull, J. J. & Antia, R. (2022). Which ‘imperfect vaccines’ encourage the evolution of higher virulence? Evolution, Medicine and Public Health, 10, 202–213.

Bull, J. J. & Lauring, A. S. (2014). Theory and empiricism in virulence evolution. PLoS Pathogens, 10, e1004387.

CDC (2026). Our history - our story. URL https://www.cdc.gov/museum/history/our-story.html.

Ch’ng, J.-H., Moll, K., Wyss, K., Hammar, U., Rydén, M., Kämpe, O., Färnert, A. & Wahlgren, M. (2021). Enhanced virulence of Plasmodium falciparum in blood of diabetic patients. PLOS One, 16, e0249666.

Collins, W. E. & Jeffery, G. M. (1999). A retrospective examination of sporozoite- and trophozoite-induced infections with Plasmodium falciparum: development of parasitologic and clinical immunity during primary infection. Am. J. Trop. Med. Hyg., 61, 4–19.

Cunnington, A. J., Riley, E. M. & Walther, M. (2013). Stuck in a rut? Reconsidering the role of parasite sequestration in severe malaria syndromes. Trends in Parasitology, 29, 585–592.

de Montjoie Rudolf, G. (1927). Therapeutic malaria. Humphrey Milford, Oxford University Press.

Eichner, M., Diebner, H. H., Molineaux, L., Collins, W. E., Jeffery, G. M. & Dietz, K. (2001). Genesis, sequestration and survival of Plasmodium falciparum gametocytes: Parameter estimates from fitting a model to malariatherapy data. Transactions of the Royal Society of Tropical Medicine and Hygiene, 95, 497–501.

Färnert, A., Snounou, G., Rooth, I. & Björkman, A. (1997). Daily dynamics of Plasmodium falciparum subpopulations in asymptomatic children in a holoendemic area. American Journal of Tropical Medicine and Hygiene, 56, 538–547.

Frank, S. (1996). Models of parasite virulence. Quarterly Review of Biology, 71, 37–78.

Fraser, C., Lythgoe, K., Leventhal, G. E., Shirreff, G., Hollingsworth, T. D., Alizon, S. & Bonhoeffer, S. (2014). Virulence and pathogenesis of HIV-1 infection: an evolutionary perspective. Science, 343, 1243727.

Freitas-Junior, L. H., Bottius, E., Pirrit, L. A., Deitsch, K. W., Scheidig, C., Guinet, F., Nehrbass, U., Wellems, T. E. & Scherf, A. (2000). Frequent ectopic recombination of virulence factor genes in telomeric chromosome clusters of P. falciparum. Nature, 407, 1018–1022.

Gandon, S., Mackinnon, M. J., Nee, S. & Read, A. F. (2001). Imperfect vaccines and the evolution of pathogen virulence. Nature, 414, 751–756.

Garnham, P. C. C. (1966). Malaria Parasites And Other Haemosporidia. 1st edn. Blackwell Scientific Publications, Oxford.

Gnangnon, B., Duraisingh, M. T. & Buckee, C. O. (2021). Deconstructing the parasite multiplication rate of Plasmodium falciparum. Trends in Parasitology, 37, 922–932.

Greischar, M. A., Beck-Johnson, L. M. & Mideo, N. (2019a). Partitioning the influence of ecology across scales on parasite evolution. Evolution.

Greischar, M. A. & Childs, L. M. (2023). Extraordinary parasite multiplication rates in human malaria infections. Trends in Parasitology, 39, 626–637.

Greischar, M. A. & Childs, L. M. (2025). Developmental synchrony and extraordinary multiplication rates in pathogenic organisms. Philosophical Transactions of the Royal Society B: Biological Sciences, 380, 20230337.

Greischar, M. A., Mideo, N., Read, A. F. & Bjørnstad, O. N. (2016a). Predicting optimal transmission investment in malaria parasites. Evolution, 70, 1542–1558.

Greischar, M. A., Mideo, N., Read, A. F. & Bjørnstad, O. N. (2016b). Quantifying transmission investment in malaria parasites. PLOS Computational Biology, 12, e1004718.

Greischar, M. A., Reece, S. E. & Mideo, N. (2016c). The role of models in translating within-host dynamics to parasite evolution. Parasitology, 143, 905–914.

Greischar, M. A., Reece, S. E., Savill, N. J. & Mideo, N. (2019b). The challenge of quantifying synchrony in malaria parasites. Trends in Parasitology, 35, 341–355.

Greischar, M. A., Savill, N. J., Reece, S. E. & Mideo, N. (2024). How to quantify developmental synchrony in malaria parasites. Frontiers in Malaria, 2, 1386266.

Huijben, S., Nelson, W. A., Wargo, A. R., Sim, D. G., Drew, D. R. & Read, A. F. (2010). Chemotherapy, within-host ecology and the fitness of drug-resistant malaria parasites. Evolution, 64, 2952–2968.

Humphreys, M. (2003). Whose body? Which disease? Studying malaria while treating neurosyphilis. In: Useful bodies: humans in the service of medical science in the twentieth century (eds. Goodman, J., McElligott, A. & Marks, L.), chap. 3. The John Hopkins University Press, Baltimore, pp. 53–77.

Kennedy, D. A. (2023). Death is overrated: the potential role of detection in driving virulence evolution. Proceedings of the Royal Society B: Biological Sciences, 290, 20230117.

Klein, E. Y., Graham, A. L., Llinás, M. & Levin, S. (2014). Cross-reactive immune responses as primary drivers of malaria chronicity. Infection and immunity, 82, 140–51.

Kriek, N., Tilley, L., Horrocks, P., Pinches, R., Elford, B. C., Ferguson, D. J., Lingelbach, K. & Newbold, C. I. (2003). Characterization of the pathway for transport of the cytoadherence-mediating protein, pfemp1, to the host cell surface in malaria parasite-infected erythrocytes. Molecular microbiology, 50, 1215–1227.

Lensen, A., Bril, A., van de Vegte, M., van Gemert, G. J., Eling, W. & Sauerwein, R. (1999). Plasmodium falciparum: infectivity of cultured, synchronized gametocytes to mosquitoes. Experimental Parasitology, 91, 101–103.

Lindblade, K. A., Steinhardt, L., Samuels, A., Kachur, S. P. & Slutsker, L. (2013). The silent threat: Asymptomatic parasitemia and malaria transmission. Expert Review of Anti-Infective Therapy, 11, 623–639.

Liu, S., Zulawinska, J., Ebel, E. R., Luniewski, A., Danis, C., Simpson, M. L., Kim, J., Ene, N., Braukmann, T. W. A., Congdon, M., Santos, W., Yeh, E. & Guler, J. L. (2025). Direct long-read visualization reveals hidden variation in GCH1 gene copy number and precise expansion steps. BMC Genomics, 26.

Mackinnon, M. J. & Read, A. F. (1999). Genetic relationships between parasite virulence and transmission in the rodent malaria Plasmodium chabaudi. Evolution, 53, 689–703.

Mackinnon, M. J. & Read, A. F. (2004). Virulence in malaria: an evolutionary viewpoint. Philosophical transactions of the Royal Society of London. Series B, Biological sciences, 359, 965–986.

McKenzie, F. E., Smith, D. L., O’Meara, W. P. & Riley, E. M. (2008). Strain theory of malaria: the first 50 years. Advances in parasitology, 66, 1–46.

Mideo, N., Reece, S. E., Smith, A. L. & Metcalf, C. J. E. (2013). The Cinderella Syndrome: Why do malaria-infected cells burst at midnight? Trends in Parasitology, 29, 10–16.

Miller, I. F. & Metcalf, C. J. E. (2022). Assessing the risk of vaccine-driven virulence evolution in SARS-CoV-2. Royal Society Open Science, 9.

Mu, J., Awadalla, P., Duan, J., McGee, K. M., Joy, D. A., McVean, G. A. T. & Su, X.-z. (2005). Recombination hotspots and population structure in Plasmodium falciparum. PLoS Biology, 3, e335.

Pak, D., Kamiya, T. & Greischar, M. A. (2024). Proliferation in malaria parasites: How resource limitation can prevent evolution of greater virulence. Evolution, 78, 1287–1301.

Patterson, D. D., Childs, L. M., Stopard, I. J., Chitnis, N., Serrato-Arroyo, S. & Greischar, M. A. (2026). Immunity can impose a reproduction–survival tradeoff on human malaria parasites. Evolution, 80, 396–412.

Pelletier, B. C. (2025). An Ill-bred Culture of Experimentation: Malaria Therapy and Race in the United States Public Health Service Laboratory at the South Carolina State Hospital, 1932-1952. Journal of the History of Medicine and Allied Sciences, 80, 67–91.

Reilly, H. B. (2007). The genetic dissection of differential growth in Plasmodium falciparum and its relationship to chloroquine drug selection. Ph.D. thesis, Notre Dame, Indiana.

Reilly, H. B., Wang, H., Steuter, J. A., Marx, A. M. & Ferdig, M. T. (2007). Quantitative dissection of clone-specific growth rates in cultured malaria parasites. International Journal for Parasitology, 37, 1599–1607.

Saad-Roy, C. M., Grenfell, B. T., Levin, S. A., Pellis, L., Stage, H. B., Van Den Driessche, P. & Wingreen, N. S. (2021a). Superinfection and the evolution of an initial asymptomatic stage. Royal Society Open Science, 8, 202212.

Saad-Roy, C. M., Grenfell, B. T., Levin, S. A., Van Den Driessche, P. & Wingreen, N. S. (2021b). Evolution of an asymptomatic first stage of infection in a heterogeneous population. Journal of the Royal Society Interface, 18, 20210175.

Saad-Roy, C. M., Wingreen, N. S., Levin, S. A. & Grenfell, B. T. (2020). Dynamics in a simple evolutionary-epidemiological model for the evolution of an initial asymptomatic infection stage. Proceedings of the National Academy of Sciences of the United States of America, 117, 1–14.

Simpson, J. A., Aarons, L., Collins, W. E., Jeffery, G. M. & White, N. J. (2002). Population dynamics of untreated Plasmodium falciparum malaria within the adult human host during the expansion phase of the infection. Parasitology, 124, 247–263.

Smalley, M. E. & Sinden, R. E. (1977). Plasmodium falciparum gametocytes: their longevity and infectivity. Parasitology, 74, 1–8.

The World Health Organization (2025). World malaria report 2025, https://www.who.int/teams/global-malaria-programme/reports/world-malaria-report-2025. Tech. rep. URL https://www.who.int/teams/global-malaria-programme/reports/world-malaria-report-2025.

Weijer, C. (1999). Another Tuskegee? American Journal of Tropical Medicine and Hygiene, 61, 1–3.

Weiss, S. (1940). Routine practices. Medical Service of ther Peter Bent Brigham Hospital. The Hospital.

Zhang, Z. (2017). Survival analysis in the presence of competing risks. Annals of Translational Medicine, 5, 47.

