## Supplemental methods, figures and tables for "No evidence for a classic transmission-duration tradeoff in human malaria infections"

### Supplemental information

#### Model fitting details

To streamline the fitting process, we fixed the number of parasite age compartments to one of three values ($n = 96, 192, 288$) representing a variance in developmental duration of 24, 12 or 8 hours (respectively). For each $n$, we calculated the SSE between predicted and observed circulating iRBCs (again $\log_{10}$ transformed) for 1000 initial parameter values chosen from a plausible range based on Latin Hypercube Sampling. Specifically, we constrained our initial parameter values to fall between zero and one for the offset parameter (which can be transformed into the initial median age, $\phi_{med}$) and for the variance in the initial Beta distribution (which specifies the initial range of developmental ages, $\phi_{range}$). We assumed $PMR_{fitted}$ varied between zero and 32, the maximum burst size for *P. falciparum* (Garnham, 1966), and allowed the initial total number of iRBCs across all developmental ages ($T$) to vary $\pm 1 \log_{10}$ unit from the initial abundance of circulating iRBCs reported in the data to be fitted. We then selected the 100 parameter sets giving the lowest SSE values for each of the three $n$ values and used these as starting values for 300 optimizations. We retained as the best fit parameters the values giving the lowest SSE value, whether or not the optimization converged. When convergence failed, it was due to plateaus in SSE values across the fitness landscape that are difficult to anticipate and avoid in such a high dimensional parameter space. We confirmed that the initial PMR values corresponding to the lowest SSE value for which the optimization converged were nearly identical to the values associated with the lowest SSE value (regardless of convergence, Fig. S8).

#### Additional supporting analyses

*Alternate definitions of virulence.* To determine whether the risk of virulence was biased by our use of an iRBC threshold-based proxy for virulence, we also considered two other virulence proxies based on drug treatment when fever exceeded a threshold temperature—either 101°F/38.3°C or 104°F/40°C (Collins & Jeffery, 1999; Weiss, 1940). All comparisons yielded similar conclusions regardless of the virulence proxy used, so we focus our analysis of model-fitted infections on the iRBC threshold proxy for virulence, which maximizes the number of infections included since fever data are missing for some patients.

*Comparing included and excluded infections.* To determine if our inclusion criteria for model-fitting inadvertently removed infections exhibiting both fast multiplication and higher risk of acute drug treatment, we compared the proportion of infections undergoing acute treatment in the fitted dataset and in the infections excluded due to a peak prior to day 6, under the assumption that faster initial PMRs would manifest as early peaks in infection. We find no difference in the proportion of acutely treated infections in these two groups (Fig. S9). Further, for the infections included (Fig. 3), initial PMRs exhibited greater values among untreated rather than treated infections, a pattern inconsistent with the idea that infections with faster growing parasite populations are more likely to be drug treated. These results bolster our original finding: there is no evidence that faster PMRs increase the risk of acute drug treatment (virulence).

We also excluded some infections based on poor model fit to data (SSE $\leq 1$). The number of fits included saturates with choice of SSE cutoff, and a cutoff of one represents a balance between including as many fits as possible and excluding poor fits (see Fig. S10). To ensure our choice of SSE cutoff did not bias our analysis, we reran our analysis using a higher cutoff of SSE $\leq 1.6$ which included 143 infections (Fig. S11 and S12, respectively).

*Competing risk analysis.* To investigate potential for strain-specific differences in the competing risks of virulence and recovery, we first test differences unrelated to strain identity. The dataset also includes information on whether the infection was initiated through injection of infected blood or via mosquito bite, and which of two hospitals administered parasites and monitored infections (Georgia State Hospital and South Carolina State Hospital, Collins & Jeffery, 1999). We find no differences due to inoculation route using the three different virulence proxies and controlling for hospital and strain (Table S1). Lacking observable inoculation-route differences, we pool infections across inoculation route to test for hospital- and strain-specific differences. For El Limon versus McLendon (the only strain comparisons possible at both hospitals due to sample sizes), we find significant differences in the risk of recovery at both Georgia State and South Carolina State (Fig. S5A, B), while the risk of virulence was only significant at South Carolina State. Importantly, while not always significant, the rank order of strain differences in virulence depended on the hospital (Fig. S5A, B).

#### Figures

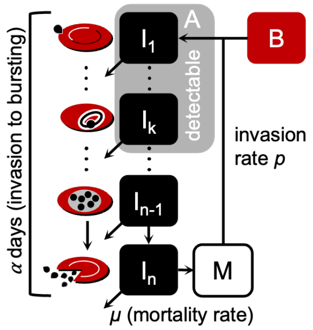


$$
\begin{aligned}
\frac{dI_1(t)}{dt} &= pB(t)M(t) - \mu I_1(t) - \frac{n}{\alpha} I_1(t) \\
\frac{dI_2(t)}{dt} &= \frac{n}{\alpha} I_1(t) - \mu I_2(t) - \frac{n}{\alpha} I_2(t) \\
&\vdots \qquad \vdots \\
\frac{dI_n(t)}{dt} &= \frac{n}{\alpha} I_{n-1}(t) - \mu I_n(t) - \frac{n}{\alpha} I_n(t) \\
A(t) &= \sum_{j=1}^{k} I_j(t)
\end{aligned}
$$

Figure S1: The data-simulating model has features that recapitulate the challenges in observed data. Within RBCs, parasites progress through $n$ developmental age classes over the $\alpha$ days required from invasion to bursting, with constant background mortality $\mu$ removing iRBCs ($I$). Of the $n$ classes, $k$ are assumed to be detectable, where $A(t)$ is detectable iRBC abundance. At the end of development, iRBCs burst to release merozoites ($M$) that invade uninfected RBCs ($B$) at rate $p$. Equations for iRBC classes on right.

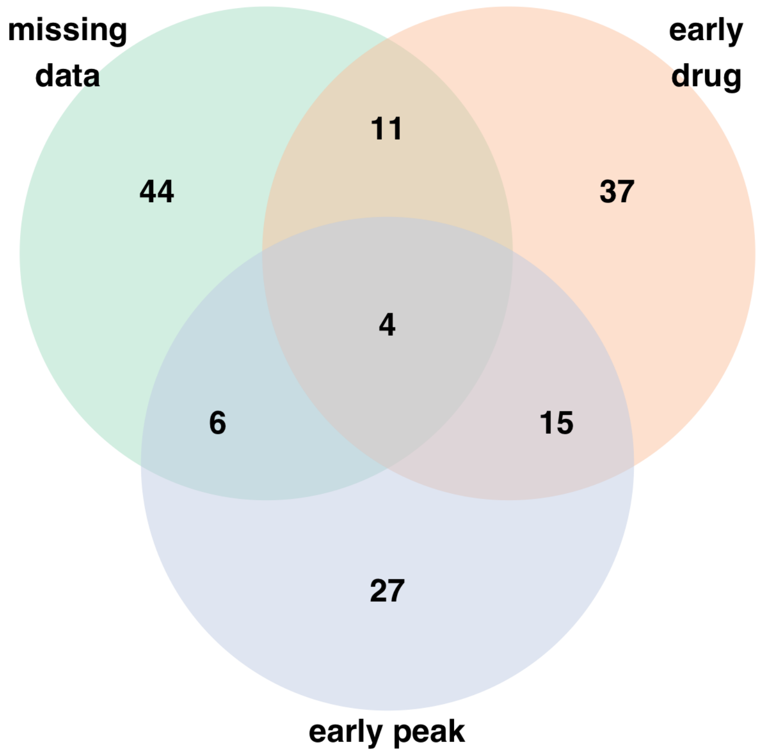


Figure S2: Reasons for excluding time series from model fitting includes infections with early peaks, with missing data in the first week, or with early drug treatment. In addition, 17 infections using strains not well-represented in the malariatherapy dataset were excluded.

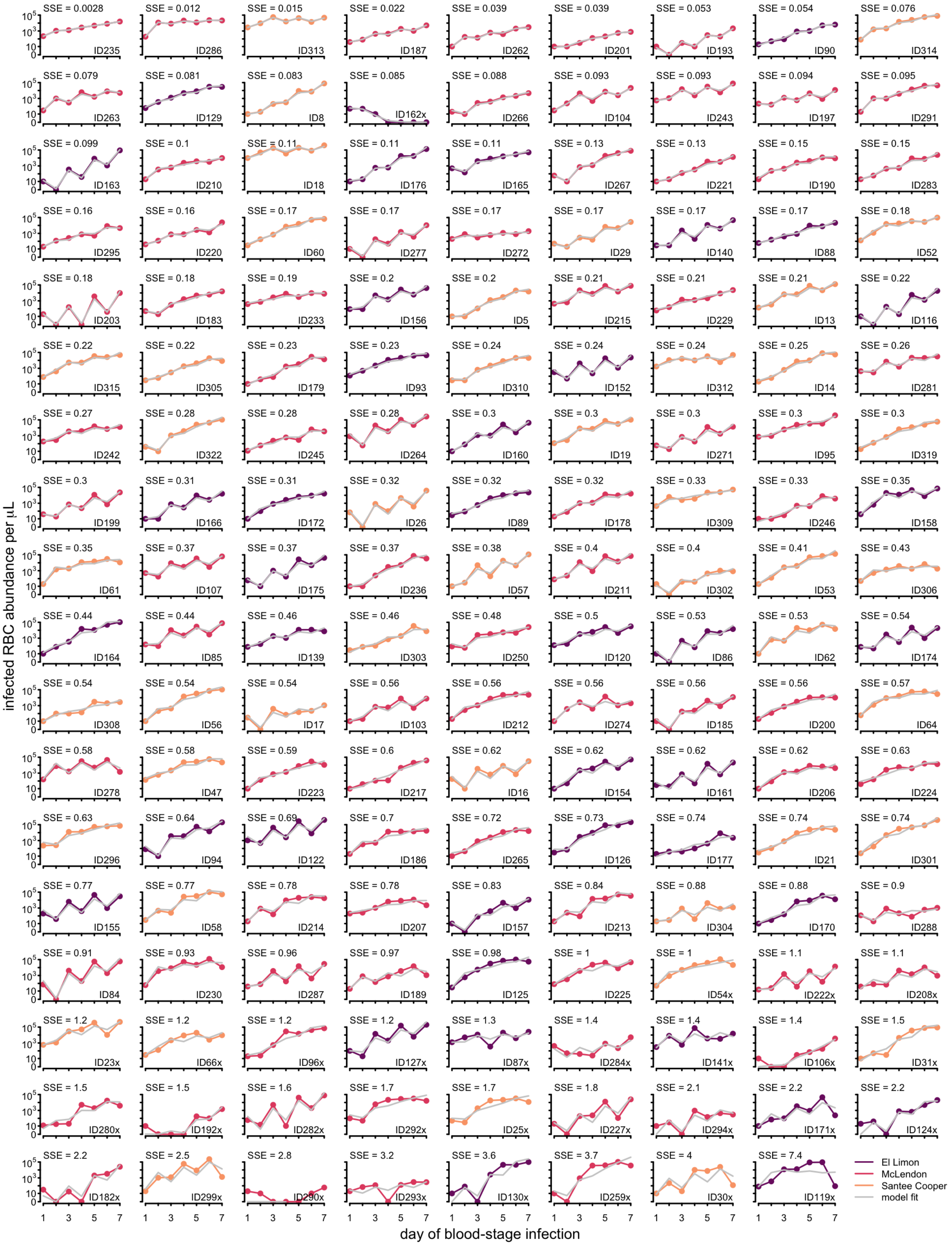


Figure S3: Time series for all 161 fitted infections ordered by increasing SSE, color coded by strain (see key at lower right) with model fit overlaid in gray. The ID of the infection is shown at lower right in each panel, followed by an ‘x’ if that fit was excluded from our main analysis.

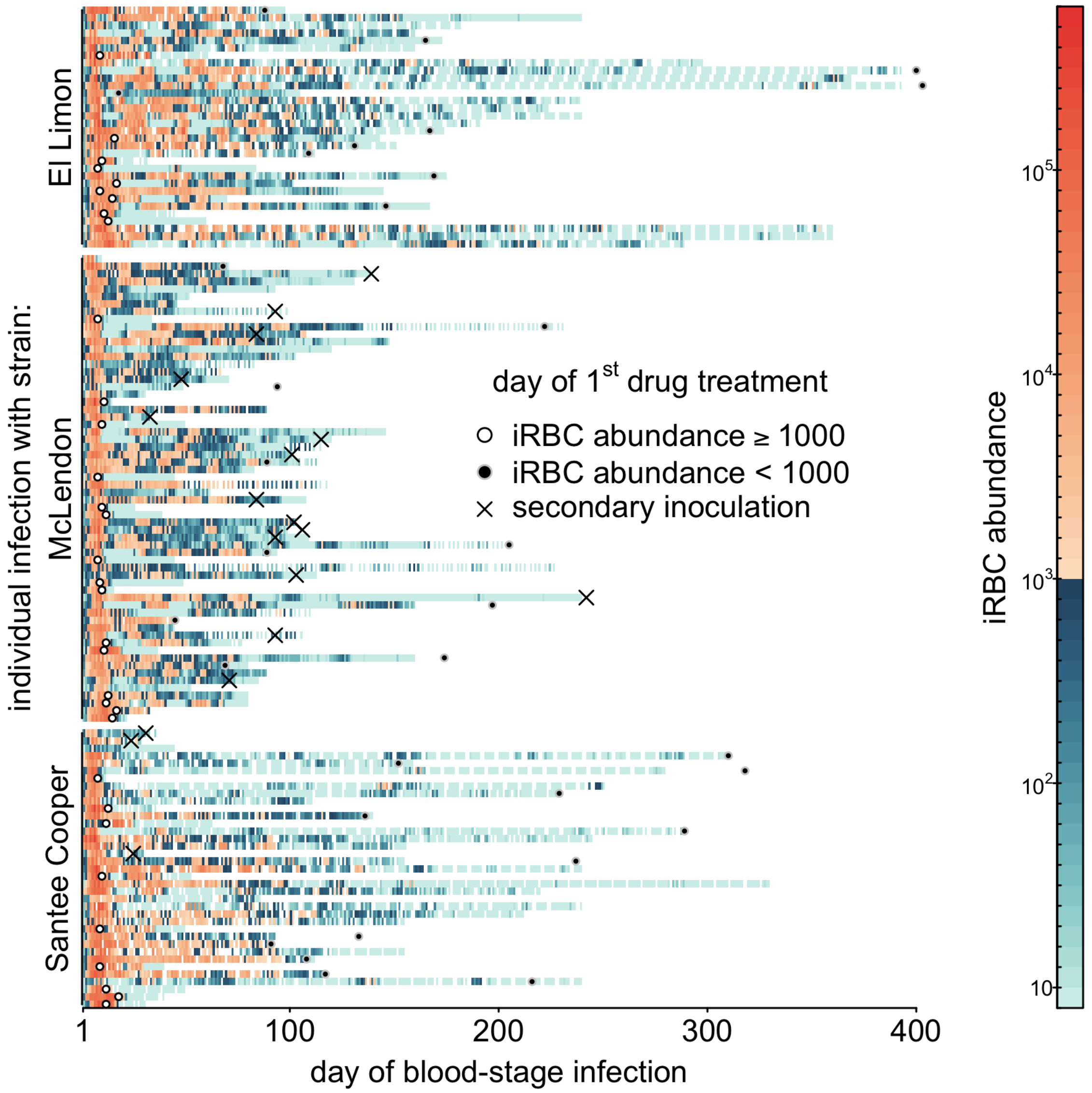


Figure S4: The abundance of iRBCs is shown as a heatmap for all infections in our analysis. Rows represent individual patients where color indicates reported iRBC abundance with circles indicating the timing of drug treatment (open for acute drug treatment, else closed), and the timing of secondary inoculation shown with an 'X'. White space indicates no data reported.

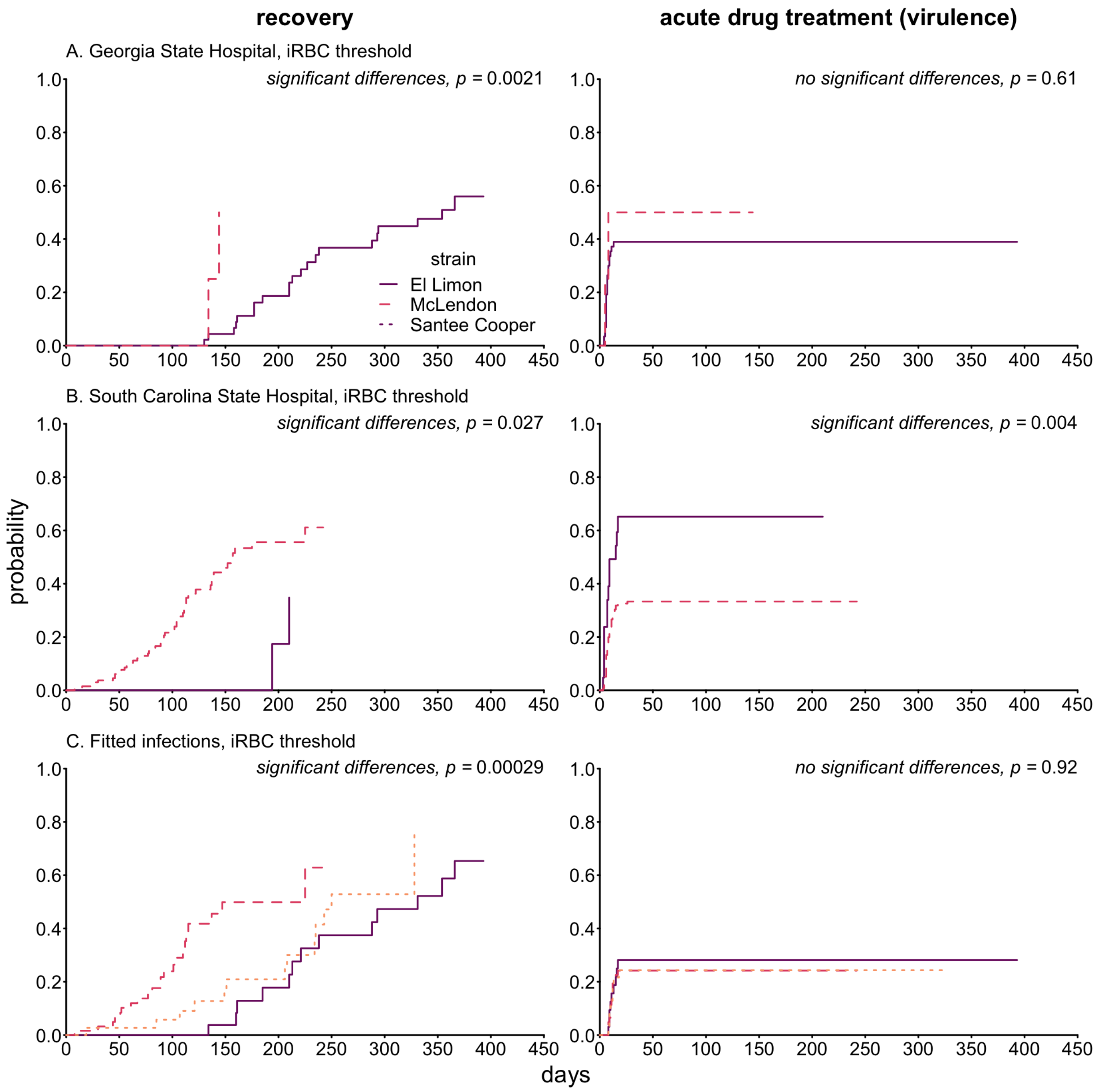


Figure S5: Competing risk analysis supports strain-specific differences in the time until recovery but inconsistent strain differences in acute drug treatment that depend on the hospital. (A) When we compare El Limon versus McLendon infections at Georgia State Hospital (full dataset), strains differ significantly in their risk of recovery (left) but not the risk of acute drug treatment (right). (B) Comparing El Limon versus McLendon infections at South Carolina State Hospital (full dataset) suggests strain-specific differences in risk of both recovery and acute drug treatment. (C) Pooling across hospital in the fitted dataset for all three strains reveals strain-specific differences in recovery but not acute drug treatment.

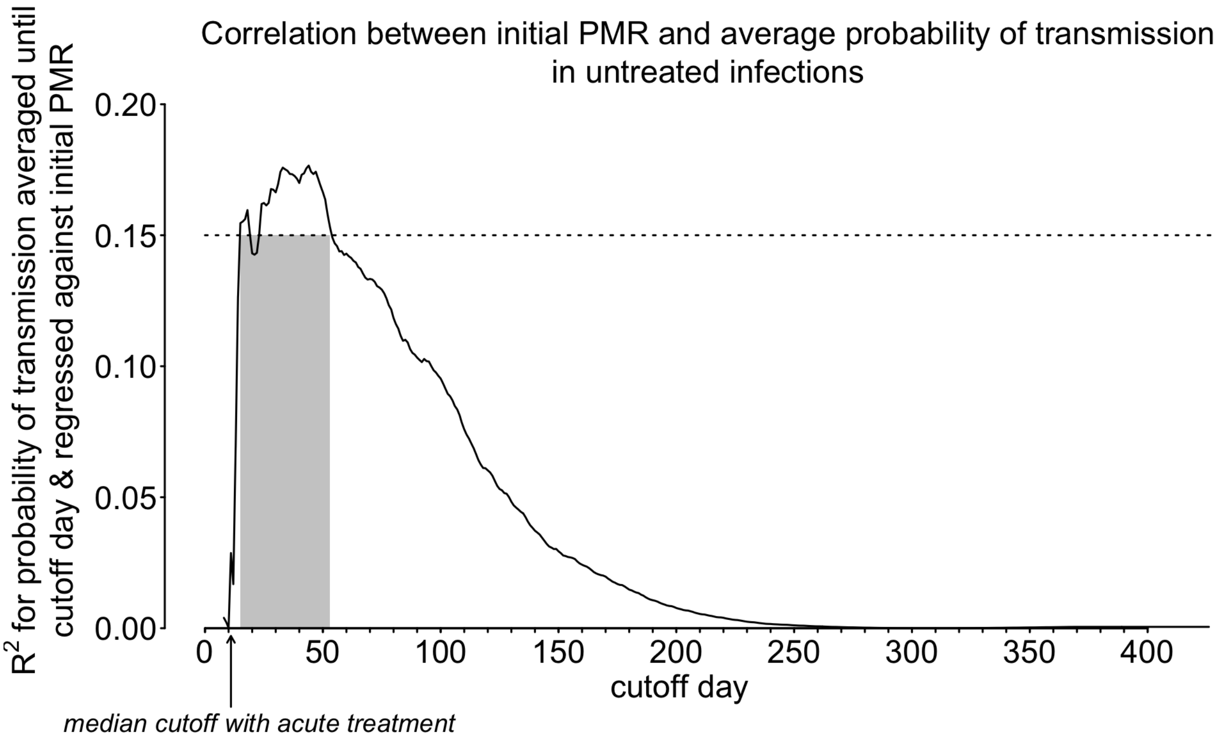


Figure S6: Initial PMRs show the greatest correlation with the probability of transmission when averaged through days 15-53 of blood-stage infection, for untreated infections (i.e., those not subject to drug treatment or secondary inoculation). To investigate the possibility of a transient increase in infectiousness due to large initial PMRs, we use data from untreated infections to re-calculate the probability of transmission averaged from the start of infection through a range of cutoff days—ranging from 8 (one day after the initial week of infection used to fit PMR values) to 427 (the last day on which transmission stages were detectable for any untreated infection). We then regress those averages against initial PMRs for each cutoff day, retaining the $R^2$ to track the strength of the correlation. Each point on the curve represents the $R^2$ obtained by regressing the average probability of transmission through the indicated cutoff day against the initial PMR. A gray rectangle indicates the first and last cutoff days with an $R^2 > 0.15$ (shown as a dotted horizontal line). For comparison with acutely treated infections, an arrow below the x-axis indicates the median time until acute treatment (11 days). Our results suggest that faster initial PMRs yield a delayed and transient boost in infectiousness, as the correlation between initial PMRs and average probability of transmission is negligible until approximately 15 days into blood-stage infection, when $R^2 > 0.15$. With a cutoff of 53 days into infection, $R^2$ declines rapidly to negligible values, i.e., the nonexistent correlation seen in Fig. 4D. For context, both *in vitro* (Lensen *et al.*, 1999) and *in vivo* (Eichner *et al.*, 2001) data suggest that *P. falciparum* requires up to 12 days to produce infectious transmission stages, and the infectivity of those stages may persist for 3 weeks (Smalley & Sinden, 1977).

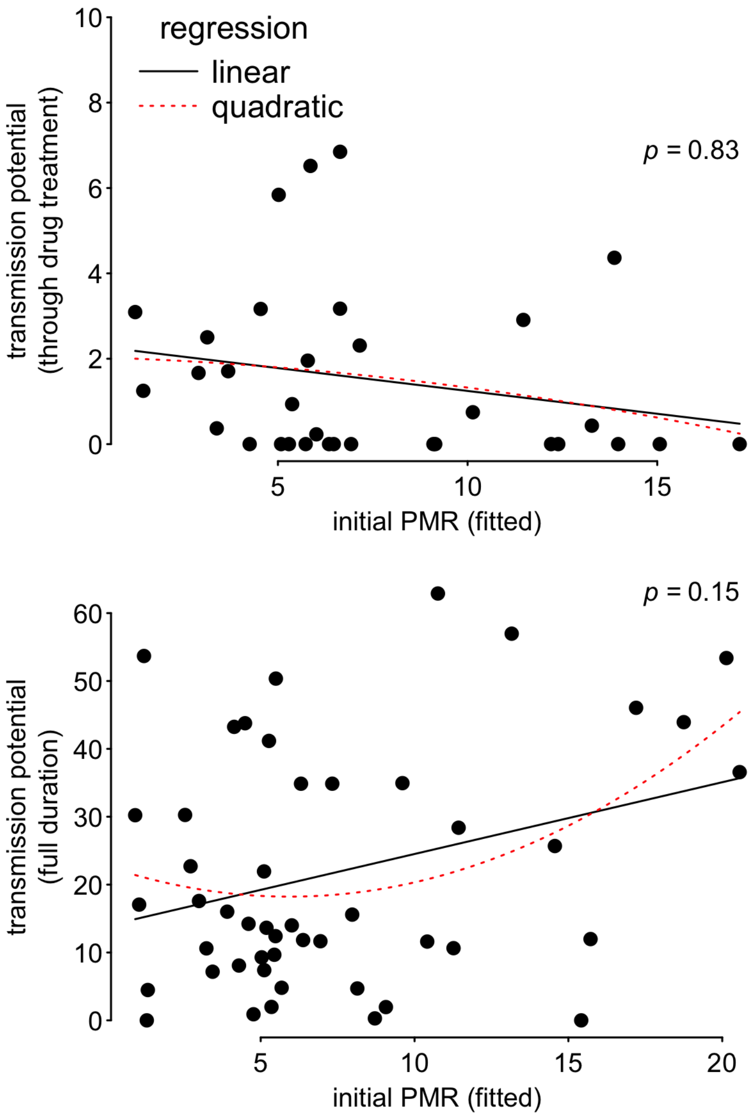


Figure S7: Quadratic regression to allow for unimodal relationships between transmission potential and $PMR_{fitted}$ gave no significant improvement over the linear regressions presented in the main text. The $p$ values associated with the ANOVA comparing quadratic and linear fits are shown in the top right of each panel.

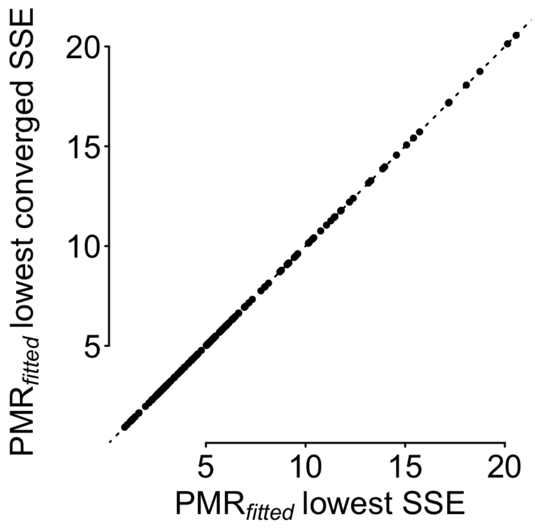


Figure S8: Fitted initial PMR values are virtually identical when comparing the value associated with the lowest SSE regardless of convergence (x-axis) and the value corresponding to the lowest SSE for which the optimization converged (y-axis).

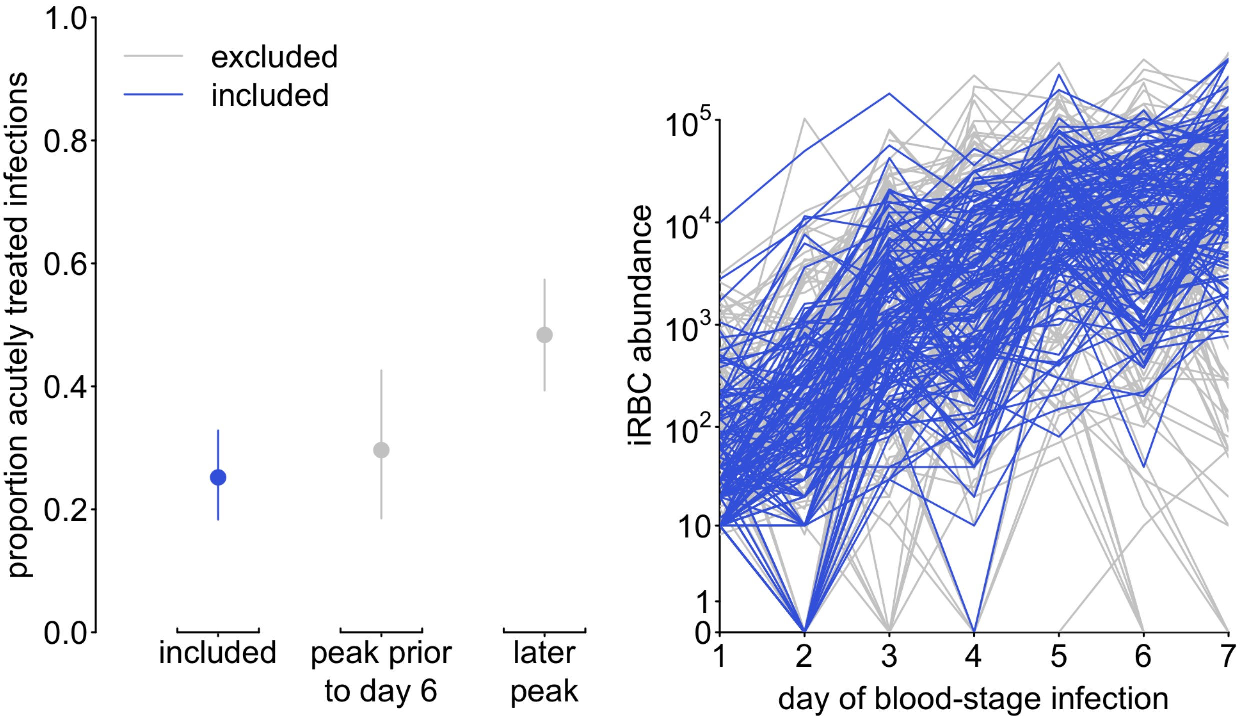


Figure S9: The proportion of acutely treated infections does not differ between included infections and excluded infections exhibiting a peak prior to day 6 (potentially due to high PMRs, left), nor do the dynamics vary much between included and excluded infections.

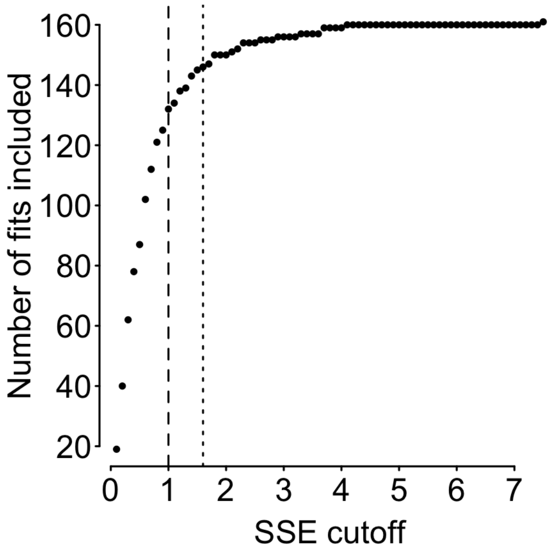


Figure S10: The number of fits that would be included (to a maximum of 161) for different SSE cutoff thresholds. Thus, the number of maximum fits includes 4 infections which were excluded due to consecutive zeros. Dashed and dotted vertical lines represent the SSE$=1$ and SSE$=1.6$ cutoffs used in our main text and supplemental analyses, respectively.

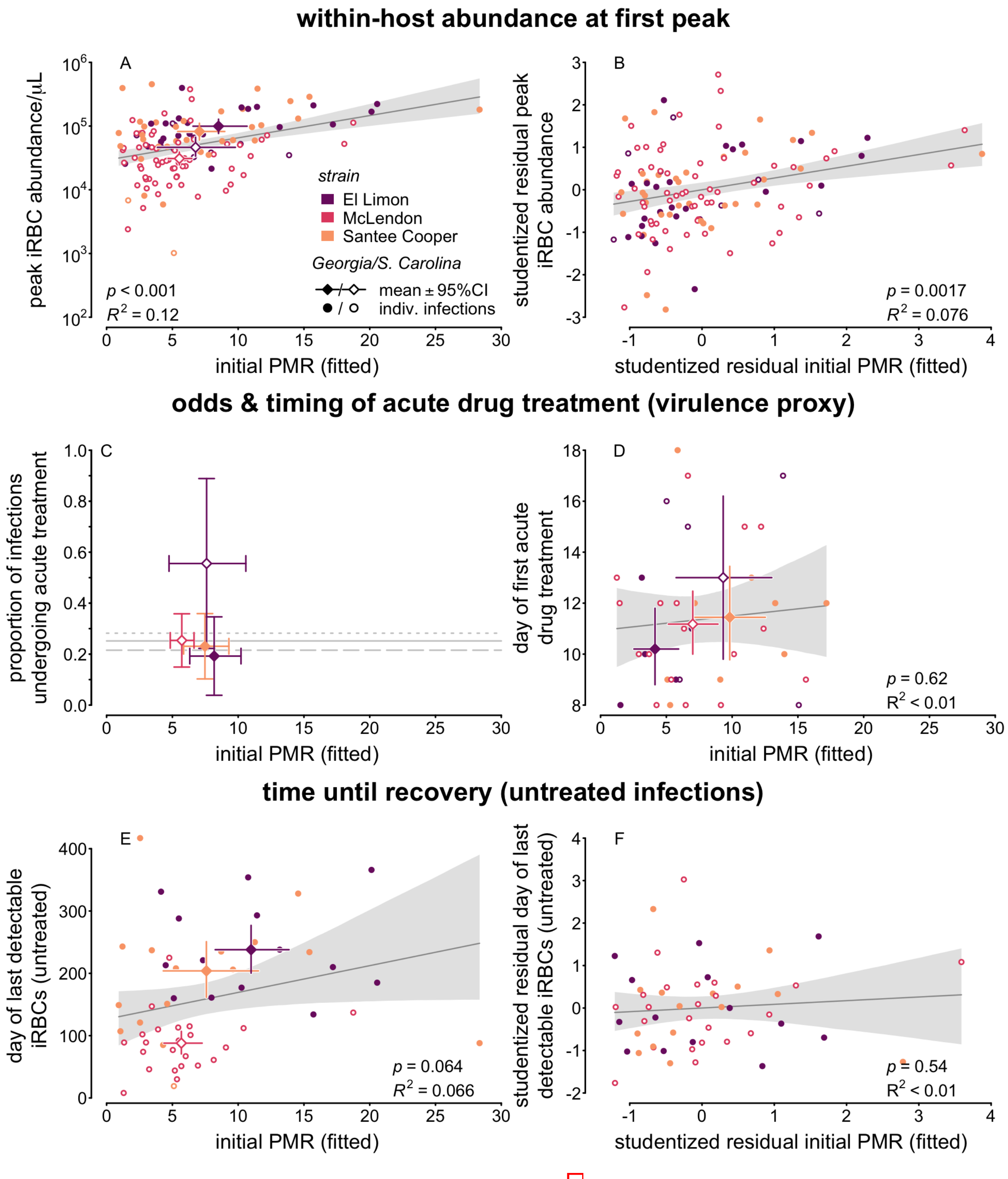


Figure S11: This figure shows the identical analyses to Fig. 3, but includes an additional 12 infections for a total sample size of 143 infections as we chose a higher cutoff of SSE$\leq$ 1.6. Linear regressions are shown as dark gray lines, with 95% confidence bounds shown as a lighter gray polygon. Qualitatively, results remain identical to the 131 included infections.

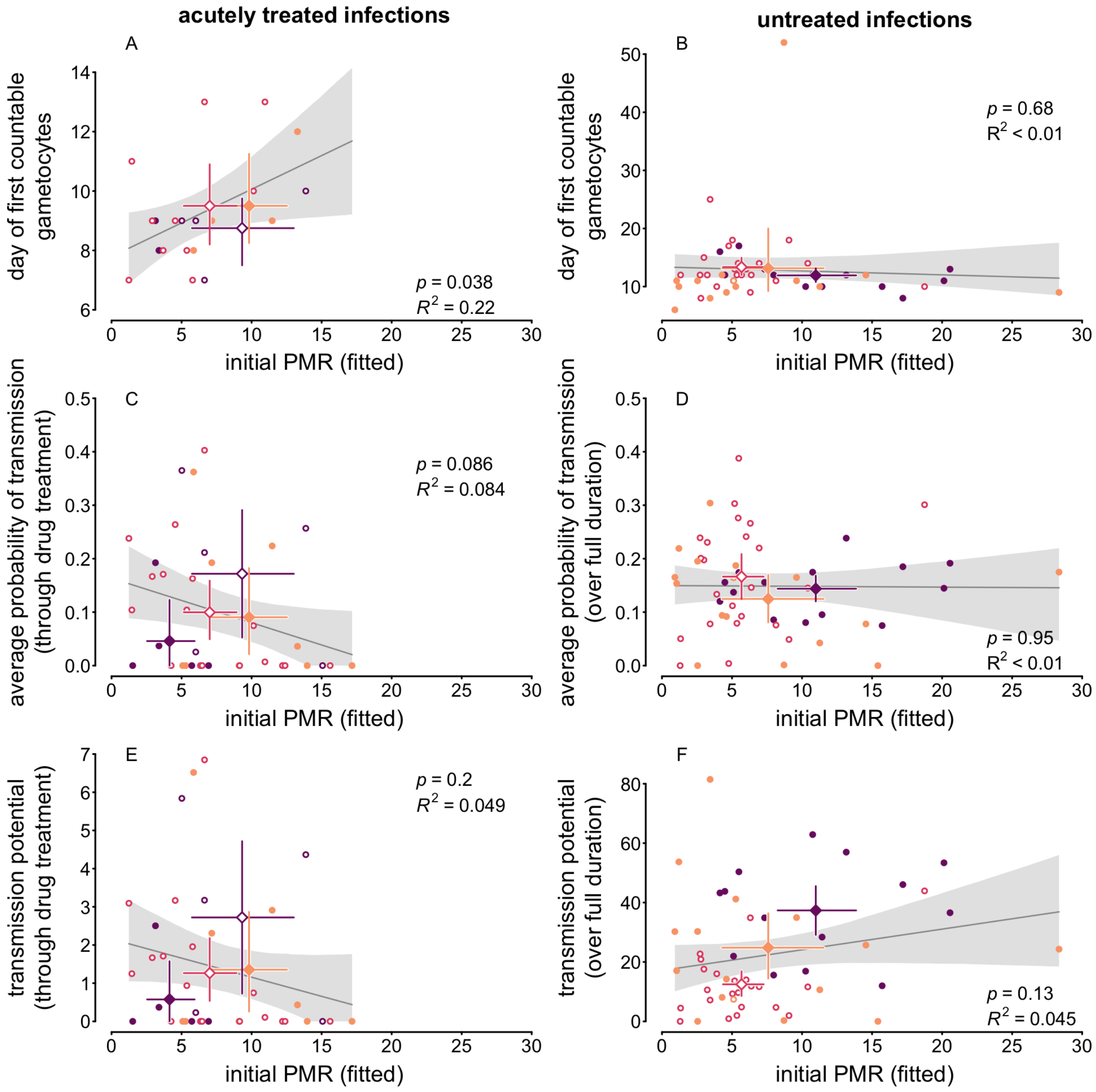


Figure S12: This figure shows the identical analyses to Fig. 4, but includes an additional 12 infections for a total sample size of 143 infections as we chose a higher cutoff of SSE$\leq$ 1.6.Linear regressions are shown as dark gray lines, with 95% confidence bounds shown as a lighter gray polygon. Qualitatively, results remain identical to the 131 included infections.

#### Tables

| | | | threshold for acute treatment (virulence proxy) | | |
|---|---|---|---|---|---|
| | | | iRBC | low fever | high fever |
| Data | | Comparison | cen./rec./vir. | cen./rec./vir. | cen./rec./vir. |
| *Inoculation route* | | | | | |
| Georgia State | El Limon | Blood vs. | 5 6 7 | 6 6 5 | 9 6 2 |
| Hospital | | Sporozoites | 10 15 15 | 8 14 14 | 12 14 10 |
| | | *recovery* | $p = 1$ | $p = 0.72$ | $p = 0.44$ |
| | | *virulence* | $p = 0.93$ | $p = 0.39$ | $p = 0.17$ |
| | McLendon | Blood vs. | 0 2 1 | 0 2 0 | 0 2 0 |
| | | Sporozoites | 0 0 1 | 1 0 0 | 1 0 0 |
| | Santee Cooper | Blood vs. | 21 24 23 | 26 19 15 | 30 19 11 |
| | | Sporozoites | 5 6 3 | 5 5 3 | 7 5 1 |
| | | *recovery* | $p = 0.33$ | $p = 0.35$ | $p = 0.5$ |
| | | *virulence* | $p = 0.34$ | $p = 0.76$ | $p = 0.32$ |
| South Carolina | El Limon | Blood vs. | 2 2 10 | 2 2 10 | 5 2 7 |
| State Hospital | | Sporozoites | 4 0 3 | 3 0 4 | 4 0 3 |
| | McLendon | Blood vs. | 32 36 29 | 35 36 24 | 39 36 20 |
| | | Sporozoites | 12 12 16 | 12 12 15 | 16 12 11 |
| | | *recovery* | $p = 0.41$ | $p = 0.32$ | $p = 0.58$ |
| | | *virulence* | $p = 0.19$ | $p = 0.076$ | $p = 0.23$ |
| | Santee Cooper | Blood vs. | 1 1 0 | 1 1 0 | 1 1 0 |
| | | Sporozoites | 0 1 0 | 0 1 0 | 0 1 0 |
| *Strain* | | | | | |
| Georgia State | | **El Limon vs.** | **15 21 22** | 14 20 19 | 21 20 12 |
| Hospital | | **McLendon** | **0 2 2** | 1 2 0 | 1 2 0 |
| | | *recovery* | $\boldsymbol{p = 0.0021}$ | | |
| | | *virulence* | $p = 0.61$ | | |
| South Carolina | | **El Limon vs.** | **6 2 13** | 5 2 14 | 9 2 10 |
| State Hospital | | **McLendon** | **44 48 45** | 47 48 39 | 55 48 31 |
| | | *recovery* | $\boldsymbol{p = 0.027}$ | $\boldsymbol{p = 0.027}$ | $\boldsymbol{p = 0.035}$ |
| | | *virulence* | $\boldsymbol{p = 0.004}$ | $\boldsymbol{p = 0.00016}$ | $\boldsymbol{p = 0.0033}$ |
| Either hospital | | El Limon vs. | 21 23 35 | 19 22 33 | 30 22 22 |
| | | McLendon vs. | 44 50 47 | 48 50 39 | 56 50 31 |
| | | Santee Cooper | 27 32 26 | 32 26 18 | 38 26 12 |
| | | *recovery* | $\boldsymbol{p = 3.2 \times 10^{-9}}$ | $\boldsymbol{p = 9.5 \times 10^{-9}}$ | $\boldsymbol{p = 3.7 \times 10^{-8}}$ |
| | | *virulence* | $p = 0.06$ | $\boldsymbol{p = 0.0071}$ | $p = 0.066$ |
| *Hospital* | | | | | |
| Either | El Limon | Georgia vs. | 15 21 22 | 14 20 19 | 21 20 12 |
| inoculation | | South Carolina | 6 2 13 | 5 2 14 | 9 2 10 |
| route | | *recovery* | $p = 0.8$ | $p = 0.99$ | $p = 0.84$ |
| | | *virulence* | $p = 0.057$ | $\boldsymbol{p = 0.013}$ | $\boldsymbol{p = 0.026}$ |

Table S1: Sample sizes for each hospital, strain, and inoculation route combination for all infections with one of the three well-represented strains (El Limon, McLendon, or Santee Cooper). *Caption continues on following page.*

Table S1: *Caption continued.* Infections were either censored ("cen.", treated for drugs outside of acute need or subject to secondary inoculation), recovered ("rec.", cleared by the patient without medical intervention), or acutely treated ("vir.", drug treated due to acute need according to one of three definitions). No patient deaths were recorded in these data, but we assume infections would have ended via disease-induced mortality (virulence) had patients not received drug treatment when in acute need (see main text). Sample sizes are greatest for the iRBC threshold proxy for virulence (drug treatment when iRBCs$\geq 1000/\mu$L), since some patients' fever data are missing. The other two proxies for virulence have lower sample sizes (drug treatment at a low fever threshold, $\geq 101°$F or drug treatment at a high fever threshold, $\geq 104°$F). Bolded combinations have sufficient sample sizes to make comparisons across inoculation route, i.e., both routes have at least one infection each in recovered and acutely treated categories, with $p$ values associated with differences in compared groups shown below for recovery and virulence (bolded $p < 0.05$). Bolded comparisons and sample sizes are shown in Fig. S5A and B.

| | | | threshold for acute treatment (virulence proxy) | | | | | | | | |
|---|---|---|---|---|---|---|---|---|---|---|---|
| | | | iRBC | | | low fever | | | high fever | | |
| Data | | Comparison | cen./rec./vir. | | | cen./rec./vir. | | | cen./rec./vir. | | |
| *Inoculation route* | | | | | | | | | | | |
| Georgia State | El Limon | Blood vs. | 3 | 2 | 2 | 3 | 2 | 2 | 4 | 2 | 1 |
| Hospital | | Sporozoites | 4 | 11 | 2 | 5 | 10 | 0 | 5 | 10 | 0 |
| | | *recovery* | $p = 0.5$ | | | | | | | | |
| | | *virulence* | $p = 0.28$ | | | | | | | | |
| | McLendon | Blood vs. | 0 | 0 | 0 | 0 | 0 | 0 | 0 | 0 | 0 |
| | | Sporozoites | 0 | 0 | 0 | 0 | 0 | 0 | 0 | 0 | 0 |
| | Santee Cooper | Blood vs. | 14 | 11 | 8 | 15 | 10 | 5 | 17 | 10 | 3 |
| | | Sporozoites | 0 | 1 | 1 | 0 | 0 | 1 | 0 | 0 | 1 |
| | | *recovery* | $p = 0.52$ | | | | | | | | |
| | | *virulence* | $p = 0.47$ | | | | | | | | |
| South Carolina | El Limon | Blood vs. | 1 | 0 | 4 | 1 | 0 | 4 | 1 | 0 | 4 |
| State Hospital | | Sporozoites | 2 | 0 | 1 | 3 | 0 | 0 | 3 | 0 | 0 |
| | McLendon | Blood vs. | 19 | 17 | 12 | 21 | 17 | 10 | 23 | 17 | 8 |
| | | Sporozoites | 6 | 5 | 3 | 6 | 5 | 3 | 7 | 5 | 2 |
| | | *recovery* | $p = 0.99$ | | | $p = 0.89$ | | | $p = 0.96$ | | |
| | | *virulence* | $p = 0.88$ | | | $p = 0.89$ | | | $p = 0.91$ | | |
| | Santee Cooper | Blood vs. | 1 | 1 | 0 | 1 | 1 | 0 | 1 | 1 | 0 |
| | | Sporozoites | 0 | 0 | 0 | 0 | 0 | 0 | 0 | 0 | 0 |
| *Strain* | | | | | | | | | | | |
| Georgia State | | El Limon vs. | 7 | 13 | 4 | 8 | 12 | 2 | 9 | 12 | 1 |
| Hospital | | McLendon | 0 | 0 | 0 | 0 | 0 | 0 | 0 | 0 | 0 |
| South Carolina | | El Limon vs. | 3 | 0 | 5 | 4 | 0 | 4 | 4 | 0 | 4 |
| State Hospital | | McLendon | 25 | 22 | 15 | 27 | 22 | 13 | 30 | 22 | 10 |
| Either hospital | | **El Limon vs.** | **10** | **13** | **9** | 12 | 12 | 6 | 13 | 12 | 5 |
| | | **McLendon vs.** | **25** | **22** | **15** | 27 | 22 | 13 | 30 | 22 | 10 |
| | | **Santee Cooper** | **15** | **13** | **9** | 16 | 11 | 6 | 18 | 11 | 4 |
| | | *recovery* | $\boldsymbol{p = 0.00029}$ | | | $\boldsymbol{p = 0.0013}$ | | | $\boldsymbol{p = 0.00092}$ | | |
| | | *virulence* | $p = 0.92$ | | | $p = 0.95$ | | | $p = 0.85$ | | |
| *Hospital* | | | | | | | | | | | |
| Either | El Limon | Georgia vs. | 7 | 13 | 4 | 8 | 12 | 2 | 9 | 12 | 1 |
| inoculation | | South Carolina | 3 | 0 | 5 | 4 | 0 | 4 | 4 | 0 | 4 |

Table S2: Sample sizes for each hospital, strain, and inoculation route combination for fitted infections. The bolded multistrain comparison is shown in Fig. S5C.
